# Rare variants and survival of patients with idiopathic pulmonary fibrosis

**DOI:** 10.1101/2024.10.12.24315151

**Authors:** Aitana Alonso-Gonzalez, David Jáspez, José M. Lorenzo-Salazar, Shwu-Fan Ma, Emma Strickland, Josyf Mychaleckyj, John S. Kim, Yong Huang, Ayodeji Adegunsoye, Justin M. Oldham, Iain Steward, Philip L. Molyneaux, Toby M. Maher, Louise V. Wain, Richard J. Allen, R. Gisli Jenkins, Jonathan A. Kropski, Brian Yaspan, Timothy S. Blackwell, David Zhang, Christine Kim Garcia, Fernando J. Martinez, Imre Noth, Carlos Flores

## Abstract

**Background:** The clinical course of idiopathic pulmonary fibrosis (IPF) is highly variable and unpredictable, with multiple genetic variants influencing IPF outcomes. Notably, rare pathogenic variants in telomere-related genes are associated with poorer clinical outcomes in these patients. Here we assessed whether rare qualifying variants (QVs) in monogenic adult-onset pulmonary fibrosis (PF) genes are associated with IPF survival. Using polygenic risk scores (PRS), we also evaluated the influence of common IPF risk variants in individuals carrying these QVs.

**Methods:** We identified QVs in telomere and non-telomere genes linked to monogenic PF forms using whole-genome sequences (WGS) from 888 Pulmonary Fibrosis Foundation Patient Registry (PFFPR) individuals. We also derived a PRS for IPF (PRS-IPF) from 19 previously published common sentinel IPF variants. Using regression models, we then examined the mutual relationships of QVs and PRS-IPF and their association with survival. Validation of results was sought in WGS from an independent IPF study (PROFILE, n=472), and results from the two cohorts were meta-analyzed.

**Results:** Carriers of QVs in monogenic adult-onset PF genes, representing nearly 1 out of 6 IPF patients, were associated with lower PRS-IPF (Odds Ratio [OR]: 1.79; 95% Confidence Interval [CI]: 1.15-2.81; p=0.010) and shorter survival (Hazard Ratio [HR]: 1.53; 95% CI: 1.12-2.10; p=7.3×10^-3^). Notably, carriers of pathogenic variants at telomere genes showed the strongest association with survival (HR: 1.76; 95% CI: 1.13-2.76; p=0.013). The meta-analysis of the results showed a consistent direction of effect across both cohorts.

**Conclusions:** We revealed the opposite effects of QVs and PRS-IPF on IPF survival. Thus, a distinct IPF molecular subtype might be defined by QVs in monogenic adult-onset PF genes. Assessing the carrier status for QVs and modelling PRS-IPF promises to further contribute to predicting disease progression among IPF patients.

## INTRODUCTION

Idiopathic pulmonary fibrosis (IPF) is a rare and progressive disease characterized by lung scarring and poor prognosis, with a median survival of 3-5 years after diagnosis (1). The clinical course of the disease varies greatly among patients and is difficult to predict. While some patients maintain relatively stable lung function for many years, 10-15% of them experience a rapid decline (2) and may succumb to their disease before initiation of effective therapy or lung transplantation. Identifying those in need of immediate therapy is crucial to improve clinical management.

Genetic studies have revealed that both rare and common genetic variants contribute to IPF susceptibility (3–8). However, the incorporation of genetic data in IPF diagnosis remains limited, as current guidelines rely primarily on radiological or histological criteria to identify interstitial pneumonia patterns. Instead, genetic testing is increasingly valuable for predicting disease prognosis (9). Genome-wide association studies (GWAS) have revealed multiple common loci involved in IPF progression and survival. For instance, the mucin 5B, oligomeric mucus/gel-forming gene (*MUC5B)* risk allele (rs3570950-T), which is the strongest common genetic risk factor known for IPF, is also linked to slower disease progression (10), although this association might be subject to an index event bias (11). A novel genetic risk locus involving the antisense RNA gene of protein kinase N2 (*PKN2*) gene has also shown association with forced vital capacity (FVC) decline, a common measure for monitoring disease progression (12). In addition, the first GWAS of IPF survival found a variant in *PCSK6* associated with differential patient survival (13).

Rare qualifying variants (QVs) in telomere-related genes were associated with poor clinical outcomes among IPF patients (7) and among patients with other interstitial lung diseases (ILD), such as hypersensitivity pneumonitis (14). Specifically, QVs associate with progressive PF, a rapid decline in lung function, and reduced survival (15–17). However, previous studies have focused mostly on the effects of a few genes (*TERT, RTEL1*, and *PARN*), even though many other telomere genes are known to be associated with monogenic PF.

Altogether, this evidence supports that multiple genetic factors are involved in distinct mechanisms of IPF pathogenesis and rates of progression. In addition, patients with the *MUC5B* risk allele are less likely to carry rare likely deleterious variants in adult-onset PF genes than non-carriers of the *MUC5B* risk allele, suggesting that the expectation of additive effects of common and rare variants may not hold in this case (7,18). Nevertheless, the effect of QVs across all known monogenic adult-onset PF genes (including telomere and non-telomere genes) in survival and the modifier role of polygenic risk scores (PRS) of common risk variants of IPF (PRS-IPF) in QVs carriers remains to be elucidated.

Using whole-genome sequencing (WGS) from IPF patients, we aimed to determine the prevalence of QVs in monogenic adult-onset PF genes and examine the mutual relationships of QVs and PRS-IPF and their association with IPF survival.

## METHODS

### Study design and sample description

We assessed the association of QVs and PRS-IPF with the primary outcome in patients with IPF. In the discovery stage, we utilized data from the Pulmonary Fibrosis Foundation Patient Registry (PFFPR) (19). In the second stage, we employed data from the Prospective Observation of Fibrosis in the Lung Clinical Endpoints (PROFILE) cohort for validation (20,21), ensuring the robustness of our findings. In the PFFPR, the primary outcome was the time from initial diagnosis to either death or lung transplantation. In the PROFILE cohort, the primary outcome was the time from diagnosis to death. For both cohorts, right censoring was applied at 60 months.

In the validation stage, we included 888 patients clinically diagnosed with IPF from the PFFPR, with baseline and longitudinal demographic and clinical information recorded in the United States since March 2016. In the second stage, 472 clinically diagnosed IPF patients from the PROFILE cohort, recruited in UK from 2010 to 2017, were included and followed for three years to track disease progression **(Figure 1).** For further details, see **Supplementary methods**, and **Supplementary Table 1.**

**Figure 1.**
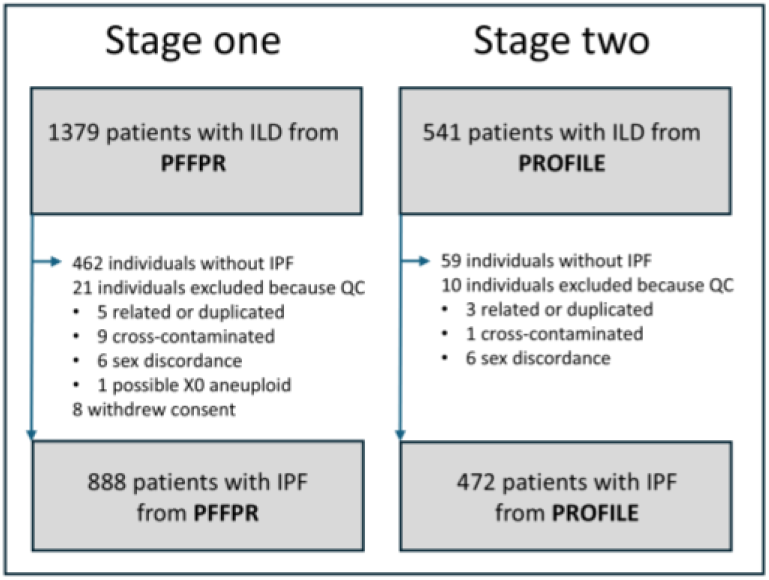
Patient cohorts included from the PFFPR and PROFILE studies.

Both studies were conducted according to The Code of Ethics of the World Medical Association (Declaration of Helsinki) and written informed consent was obtained from all participants. The Research Ethics Committees at each participating centre approved the study.

### Sequencing and bioinformatics analysis methods

In the PFFPR, library preparation and sequencing were performed by Psomagen (Rockville, MD). Genomic DNA libraries were prepared using the TruSeq DNA PCR Free kit (Illumina Inc.) and sequenced on an Illumina NovaSeq 6000 instrument (Illumina Inc.) with 150 bp paired-end reads at an average depth of 30X. At least 80% of the genome was covered by ≥20 reads, and ≥90% was covered by ≥10 reads. WGS was processed using the Illumina DRAGEN Bio-IT Platform Germline Pipeline v3.10.4 (Illumina Inc.) using the Illumina DRAGEN Multigenome Graph hg38 as the reference genome. Only variants with a “PASS” filter were included in subsequent analyses.

For the PROFILE cohort, WGS was performed at Human Longevity Inc. using the Illumina NovaSeq 6000 system with 150 pb paired-end reads. Coverage of at least 10X was achieved in over 98% of the Consensus Coding Sequence Release 22 (CCDS), with an average read depth of 42X across the CCDS as described previously (4). Sequences were processed using the Illumina DRAGEN Bio-IT Platform Germline Pipeline v3.0.7, with the GRCh38 as the reference genome.

In both cohorts, quality control (QC) included identifying QC outliers, detecting kinship between patients, checking for cross-contamination of samples, and identifying sex discordance, using metrics from different tools. **Figure 1** summarized the number of individuals excluded and the reasons for exclusion. For further details, see **Supplementary methods**.

### Identification of QVs in monogenic adult-onset PF genes

We restricted the identification of QVs to a curated list of 13 PF genes, categorized as either telomere related (*TERC, TERT, TINF2, DKC1, RTEL1, PARN, NAF1*, and *ZCCHC8*) or non-telomere related (*SFTPC, SFTPA1, SFTPA2, SPDL1*, and *KIF15*) **(Supplementary Table 2)**. With the exception of *SPDL1*, and *KIF15*, this list includes genes with a known dominant inheritance pattern (presuming that QVs in these genes would have higher penetrance) and genes commonly found in familial IPF cohorts, despite they also occur in sporadic cases (7).

*KIF15* and *SPDL1* were incorporated to the list as recent largescale sequencing studies identified them as PF-related genes (4,22,23). Both genes are critical for mitosis, pointing to a novel, non-telomeric mechanism underlying IPF. Rare deleterious variants in *KIF15* and three telomere genes (*TERT, PARN* and *RTEL1*) have been previously associated with IPF risk, early onset, and progression to early-age lung transplantation or death (23). In *SPDL1,* a rare missense variant was confirmed as a new IPF risk allele, although carriers did not exhibit distinct clinical features (4). For simplicity, we refer to this gene set as monogenic adult-onset PF genes.

Variants in these genes were filtered based on read depth (DP) <10, mapping quality (MQ) <50, or the percentage of missing genotypes (FMISS) >0.05 in the cohort. The remaining variants were annotated using the Variant Effect Predictor tool v109.3 (24). Variants with a global allele frequency (AF) >0.0005 in gnomAD v2.1 were excluded from the study. For our analyses, we retained protein-truncating variants (including frameshift, stop-gained, start-loss, and splicing variants) and missense variants with a CADD >15.

For the non-coding RNA gene *TERC*, due to the difficulty in predicting functional effects in non-coding genes, variants were considered for the analysis if their global population AF was <0.0005 and they were annotated by ClinVar as pathogenic (P), likely pathogenic (LP), or of uncertain significance (VUS).

All variants that met the aforementioned criteria were annotated as the total set of QVs. Additionally, QVs were manually classified according to ACMG guidelines (25) as P, LP, or VUS. Variants classified as P or LP comprised the set of pathogenic variants, while ClinVar variants were cross-referenced and annotated as VUS, P, or LP.

For sensitivity analysis, we defined six additional categories for QVs based on specific thresholds for population AF and different predicted variant effect. We also assessed a category of rare synonymous variants, expected to capture neutral variation, to use as a null model in in the association analyses. These criteria are summarized in **Supplementary Table 3**.

### Principal components of genetic heterogeneity in the cohorts

Principal components (PC) were calculated after excluding single nucleotide polymorphisms (SNPs) with a minor allele frequency (MAF) <0.01 from WGS data, using BCFtools (https://samtools.github.io/bcftools/bcftools.html). Genotyping QC was then performed using PLINK v.1.9. First, SNPs with a genotyping call rate (CR) <95% or those deviating significantly from Hardy-Weinberg equilibrium (HWE) (p<1.0×10^-6^) were removed. After linkage disequilibrium pruning (indep-pairwise 100 5 0.01), the main PCs of genetic variation were calculated based on 110,951 independent SNPs in the PFFPR and 143,214 independent SNPs in PROFILE (see **Supplementary Figure 1)**.

### Estimation of the PRS of IPF in the cohorts

The PRS-IPF for each patient was derived using the 19 previously published genome-wide significant IPF variants **(Supplementary Table 4)** using PRSice-2 (26). Briefly, PRS were calculated as the number of risk alleles carried by each individual, multiplied by the effect size of the variant as described in the GWAS study (26) summed across all variants included in the score:

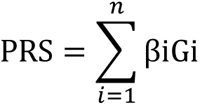

where βi is the OR (in the case of binary traits) from variant i, Gi represents the number of risk alleles carried at the variant i and n represents the conditionally independent signals identified elsewhere. Raw polygenic scores were then standardized as z-scores using the following formula:

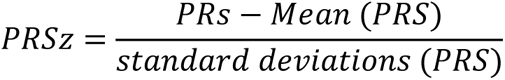

As part of the sensitivity analysis, we also used the same methodology to derive PRS for telomere length (PRS-TL) based on the 20 common variants that were previously found associated with leukocyte telomere length (TL) (27) **(Supplementary Table 5)**. In this case, since TL is a quantitative trait, βi is represented by beta coefficients in the PRS formula.

### Statistical analysis

Descriptive statistics were provided as mean (standard deviation, SD) or median (interquartile range, IQR) and valid percentage for continuous (quantitative) and categorical (binary) data, respectively. Categorical variables were compared using a Chi-squared test or a Fisher’s exact test as indicated.

To examine the relationship between the presence of QVs and the PRS, we first used the Student’s t-test and the Kolmogorov-Smirnov (KS) test to compare the mean PRS values and their distributions between QVs carriers and non-carriers. Additionally, we assessed this relationship using logistic regression models, adjusting for sex, age at diagnosis, and the two main PCs of genetic heterogeneity, which accounted for a significant proportion of genetic variance **(Supplementary Figure 1C)**.

To examine the association between QVs, PRS, and survival, we used Cox proportional hazards regression models adjusted for sex, age at diagnosis, the two main PCs for genetic heterogeneity, smoking history, DLCO% predicted, FVC % predicted, and *MUC5B* risk allele carrier status necessary. The proportional hazards assumption of each covariate was assessed by plotting scaled Schoenfeld residuals against transformed time, revealing no evidence of non-proportional hazards.

The Survival R package (v.3.5-7) was used to calculate p-values, hazard ratios (HR), and 95% confidence intervals (CI). For visualizing survival differences, we generated Kaplan-Meier survival plots and tested the differences using log-rank tests.

Results from PFFPR and PROFILE studies were meta-analysed under a fixed-effects model to assess the directional concordance of associations, using the Meta R package (28). Statistical analyses were performed with R v.4.3.1, with p-values <0.05 considered significant.

## RESULTS

### Prevalence of QVs in the PFFPR

We identified 131 QVs in monogenic adult-onset PF genes in 144 patients from the PFFPR resulting in a diagnostic yield of finding a QV of 16.2%, with a 95% CI of 13.8-18.6 **(Supplementary Figure 2**, **Table 1)**. Most patients (96.5%) carried a single QV, while five patients (3.5% of the QV carriers) had two or more QVs, including combinations such as *NAF1/TERT, KIF15/RTEL1, TERT/SPDL1, TINF2/TERT/RTEL1*, and *RTEL1/RTEL1*. Consistent with previous studies, the prevalence of QVs was higher in patients with a familial history of disease (27.3%) compared to those with sporadic disease (13.5%) (*p*=3.08×10^-5^).

**Table 1.** Count of qualifying variants in monogenic adult-onset PF genes identified in the PFFPR patients.*

| Gene | Number of variants | P/LP | VUS | Gene category |
| --- | --- | --- | --- | --- |
| *KIF15* | 15 | 3 | 12 | Non_Telomere |
| *SPDL1* | 8 | 2 | 7 | Non_Telomere |
| *SFTPC* | 5 | 0 | 5 | Non_Telomere |
| *SFTPA2* | 1 | 0 | 1 | Non_Telomere |
| *SFTPA1* | 1 | 0 | 1 | Non_Telomere |
| *ZCCHC8* | 5 | 1 | 4 | Telomere |
| *TINF2* | 3 | 3 | 0 | Telomere |
| *PARN* | 16 | 12 | 4 | Telomere |
| *RTEL1* | 33 | 18 | 15 | Telomere |
| *DKC1* | 1 | 0 | 1 | Telomere |
| *TERC* | 4 | 0 | 4 | Telomere |
| *NAF1* | 6 | 1 | 5 | Telomere |
| *TERT* | 31 | 18 | 13 | Telomere |
| **Total variants** | 131 | 58 | 73 |  |

\*Filtered by frequency (AF < 0.0005) and CADD score (>15). P/LP, Pathogenic or likely pathogenic; VUS, variant of uncertain significance.

Most QVs were in telomere genes (75.6%), while nearly a quarter were found in non-telomere genes (22.9%). The highest number of QVs were identified in telomere-related genes including *RTEL1* (25.2%), *TERT* (23.7%), and *PARN* (12.2%). These genes also had the highest proportion of P/LP variants (31.0%, 31.0%, and 20.7%, respectively) **(Table 1).** In total, 42.7% of QVs were previously annotated in ClinVar as VUS, LP, or P.

Consistent with previous findings (7,18), carriers of the risk *MUC5B* allele (rs35705950 TT or TC genotype) had lower prevalence of QVs (13.5%) compared to those carrying the protective GG genotype (22.6%) (*p*=1.03×10^-3^).

### Association of QVs and PRS-IPF in the PFFPR

Given the potential non-additive effects of QVs and the *MUC5B* common variant, we investigated whether individuals with lower polygenic risk were more likely to carry QVs compared to those with higher polygenic risks. We first compared the mean and distribution of PRS-IPF between QV carriers and non-carriers and found significant differences (Student’s t-test, *p*=1.30×10^-3^; KS-test, *p*=3.74×10^-4^) **(Figure 2A)**. When patients were stratified into PRS-IPF tertiles, the prevalence of QVs was higher in the lowest tertile patients (low PRS-IPF) than in the highest tertile patients (high PRS-IPF), which associated with an increased risk of carrying a QV in the patients classified in the low PRS-IPF tertile (OR=1.79, 95% CI=1.15-2.81, *p*=0.010) **(Figure 2B)**. The association persisted when the cohort was divided into two PRS-IPF categories, low and high (OR=1.74, 95% CI=1.20-2.53, *p*=3.57×10^-3^ **(Supplementary Figure 3)**.

**Figure 2.**
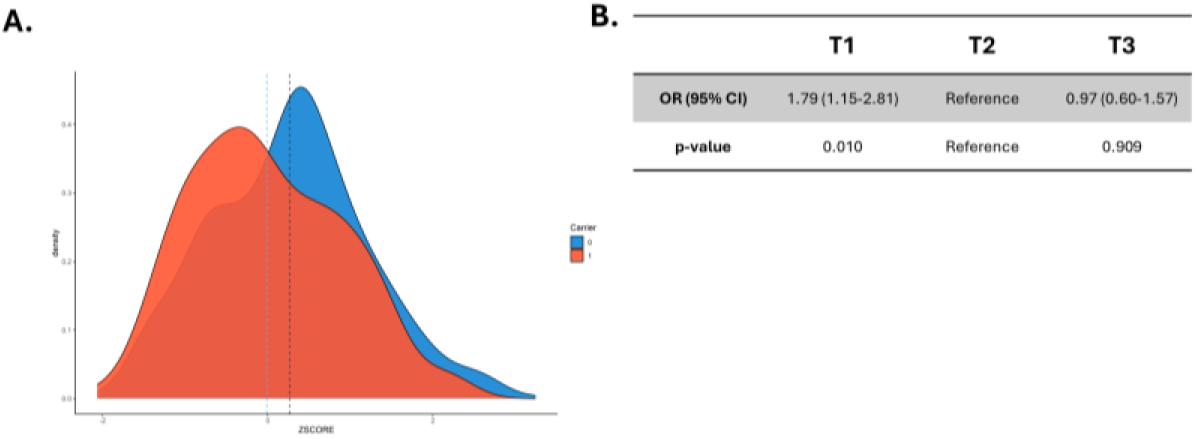
Association between prevalence of qualifying variants (QVs) and PRS-IPF in the PFFPR. A) Distribution of PRS-IPF in QV carriers (1) and non-carriers (0). Vertical dotted lines represent the mean value of the distribution. B) Risk of carrying a QV for patients with low polygenic risk (T1) and high polygenic risk (T3) compared to those in the middle tertile. The odds ratios (OR) and the 95% confidence intervals (CI) were estimated using logistic regression adjusted for sex, age of diagnosis, and the two main principal components.

Excluding the *MUC5B* locus from the PRS-IPF calculations yielded non-significant differences in the mean and distribution of PRS-IPF between QV carriers and non-carriers **(Supplementary Figure 4A)**. However, QVs remained more common in the lowest PRS tertile patients compared to the highest (OR=1.60, 95% CI=1.01-2.54, *p*=0.05) **(Supplementary Figure 4B)**.

To explore if these observations were independent of genetically predicted TL, we then assessed the association between the prevalence of QVs and PRS-TL, focusing only on the QVs in telomere genes. No significant associations were found between QVs and PRS-TL under the two assumptions **(Supplementary Figures 5-7)**.

### Association of QVs with survival in the PFFPR

Given that carriers of the *MUC5B* risk allele are associated with better survival and are less likely to carry QVs, we hypothesized that QV carriers might have poorer survival. Indeed, QVs carriers were associated with reduced survival (HR=1.53, 95% CI=1.12-2.10, *p*=7.33×10^-3^; log-rank test, p=0.022). The result was consistent when the analysis was limited to QVs that were classified as pathogenic or likely pathogenic variants (HR=1.71, 95%CI=1.11-2.65, *p*=0.016; log-rank test, p=0.043). However, no significant association was found for ClinVar variants alone (HR=1.35, 95% CI=0.87-2.09, *p*=0.18) **(Figure 3A, Supplementary Figure 8)**. As an internal control, we found no association between survival and rare synonymous variants in the same monogenic adult-onset PF genes (HR=1.38, 95%CI=0.80-2.38, *p*=0.24) **(Figure 3A)**.

**Figure 3.**
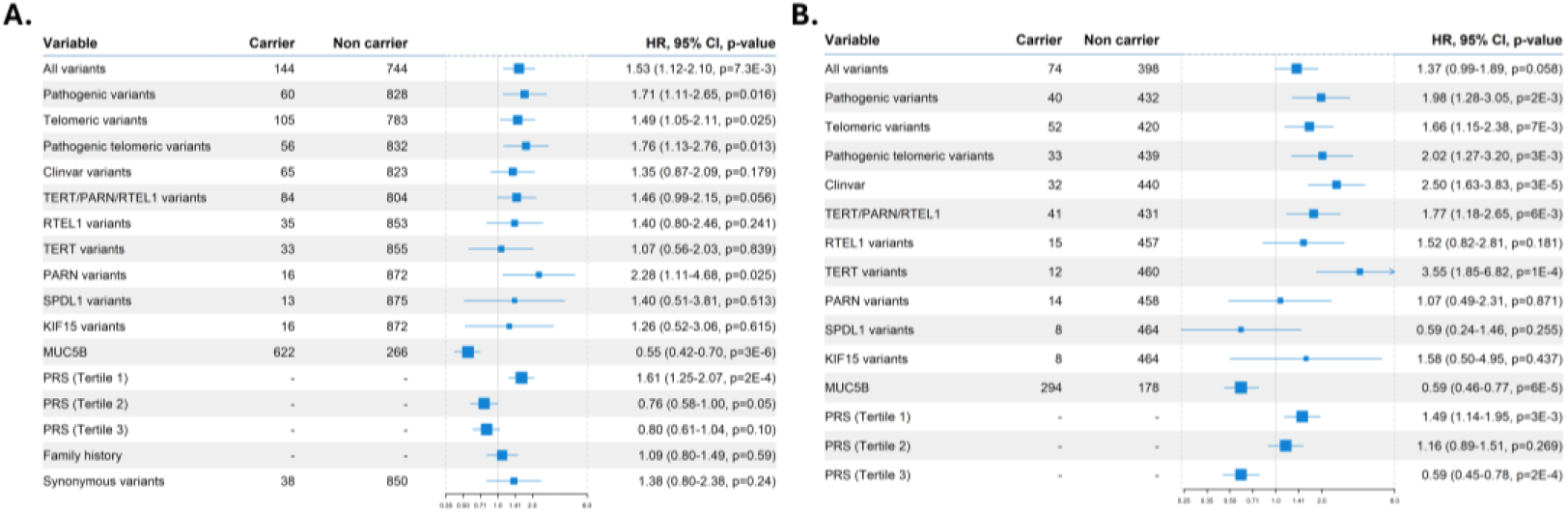
Qualifying variants (QVs), *MUC5B* risk allele, PRS-IPF, and family history effect on survival. A) PFFPR. B) PROFILE. All analysis were performed using Cox regression models adjusted for sex, age of diagnosis, the two main principal components, smoking history, forced vital capacity (FVC) % predicted, and diffusing capacity for carbon monoxide (DLCO) % predicted, and the *MUC5B* risk allele whenever necessary. The X-axis shows Hazard-ratios (HR); the grey line corresponds to the HR=1.0. The circles correspond to adjusted HR and horizontal lines correspond to 95% confidence intervals (CI).

Further analyses showed that QVs in telomere-related genes had the largest effect on survival (HR=1.76, 95%CI=1.13-2.76, *p*=0.013; log-rank test, p=0.029), and QVs in *PARN* were particularly associated with worse survival (HR=2.28, 95%CI=1.11-4.68, *p*=0.03; log-rank test, p=0.035) **(Figure 3A**, **Supplementary Figure 8)**.

We performed additional sensitivity analyses. First, excluding *PARN* QV carriers attenuated effect size, but the results remained significant (HR=1.46, 95% CI=1.04-2.03, *p*=0.03), indicating that other genes also contribute to the association with worse survival **(Supplementary Figure 9)**. Second, as the probability of carrying QVs is higher among cases with a family history of PF, and family history of PF predicts reduced survival (29), we tried to account for the risk attributed to family history of PF. We found no relationship between family history of PF and survival (HR=1.09, 95%CI=0.80-1.49, p=0.59), suggesting that family history is not a major factor in this cohort’s survival outcomes **(Figure 3A)**. Third, the criteria for defining QVs are subject to different choices and predictors. Using alternative and stricter definitions of QVs in the analyses **(Supplementary Table 3)**, we found that the effect had consistent directionality across all QV definitions **(Supplementary Figure 10)**. Additionally, we observed that applying more stringent *in silico* predictors resulted in a higher risk of reduced survival (ultrarare PTV HR=2.44, 95% CI=1.14-5.24, *p*=0.02). However, these criteria also led to a reduced number of identified carriers, thereby decreasing the power to detect significant differences.

### Associations of PRS-IPF with survival in the PFFPR

Since both rare and common IPF genetic variants are associated with IPF survival, and QVs associate with worse survival, we then examined whether the polygenic component of IPF was also associated with survival. As PRS-IPF values were mainly influenced by the *MUC5B* effect, we did not adjust for the risk *MUC5B* genotype in these analyses, although we did for relevant individual covariates. We found that the lowest PRS-IPF tertile was associated with the worst survival (log-rank test, *p*=1.8×10^-4^; HR=1.61, 95% CI=1.25-2.07, *p*=1.9×10^-4^) **(Figure 3A**, **Figure 4)**. In contrast, PRS-TL was not associated with survival, whether analyzed by tertiles or by high *vs*. low-risk groups **(Supplementary Figure 11**, **Supplementary Figure 12)**.

**Figure 4.**
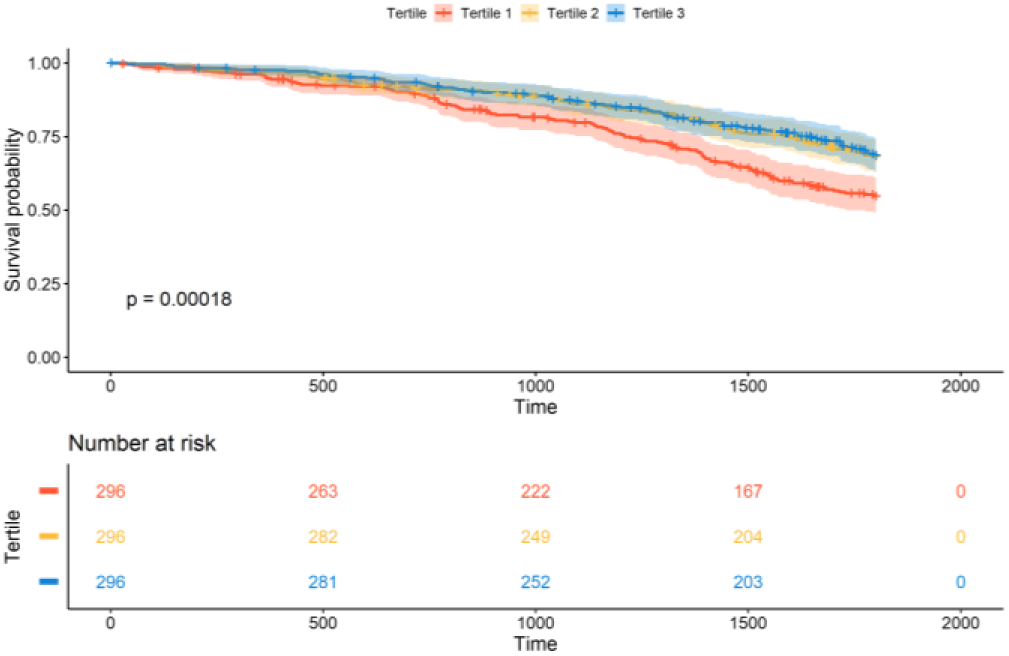
Association between PRS-IPF and survival in PFFPR. Patients with low polygenic risk of IPF (T1) have worse survival in comparison with patients with high polygenic risk of IPF (T2 and T3).

A sensitivity analysis, excluding the *MUC5B* locus from the PRS-IPF calculation, yielded non-significant results **(Supplementary Figure 13)**. To explore whether the association of PRS-IPF and survival was solely driven by the known association of *MUC5B*, we stratified patients by QV carrier status and assessed its effect in each group. The analyses showed that patients with lower PRS-IPF were associated with worse survival in both groups, although this association was attenuated among carriers (HR=1.76, 95% CI=0.99-3.15, p=0.055) compared to non-carriers (HR=1.54, 95% CI=1.16-2.04, p=2.5×10^-3^) **(Supplementary Figure 14A, Supplementary Figure 14B)**. Similar results were obtained when we assessed the effect of the *MUC5B* rs35705950 genotypes on survival among both groups **(Supplementary Figure 14C, Supplementary Figure 14D)**. Our findings suggest that the strong observed association between PRS-IPF and survival is mainly driven by *MUC5B*. While it is suspected that QVs may exert an independent but opposite effect on IPF survival, we cannot rule out that the observed differences were influenced by disparities in the group sizes.

### Validation of results in PROFILE and meta-analysis

Using the same classification of QVs as in PFFPR, the diagnostic yield of finding a QVs in PROFILE was 15.67 % (95% CI=12.4-19.0%). However, PROFILE IPF patients showed no statistical differences in the mean and the distribution of PRS-IPF between QV carriers and non-carriers (Student’s t-test, *p*=0.17; KS-test, *p*=0.24). Similarly, although not statistically significant, an enrichment of QVs in the patients with lower polygenic component of IPF was observed compared to those with higher polygenic component (OR=1.29, 95% CI=0.78-2.15, *p*=0.31) **(Supplementary Figure 15)**.

Despite these results, the association of QVs and IPF survival were validated in PROFILE patients **(Figure 3B)**. As for PFFPR, the survival analyses of PROFILE patients focusing on the LP/P variants, both of all genes or only of telomeric genes, also showed the largest effect sizes (HR=1.98, 95% CI=1.28-3.05, *p*=2.1×10^-3^; log-rank test, p=0.023). However, we did not replicate the association of QVs in *PARN* with survival. Instead, the association with worse survival in PROFILE was observed for *TERT* QV carriers (HR=3.55, 95% CI=1.85-6.82, *p*=1.4×10^-4^; log-rank test, p=0.03) **(Figure 3B**, **Supplementary Figure 16)**.

The meta-analysis of results from PFFPR and PROFILE cohorts showed a consistent direction of effect across all categories and supported a robust association between QVs (including “all variants”, “pathogenic”, “telomeric”, and “pathogenic telomeric”) and PRS-IPF tertiles with survival **(Figure 5)**. In the meta-analysis, *RTEL1* QVs were also nominally associated with IPF survival despite not reaching nominal significance in any of the two cohorts by separate. No association with IPF survival was found for QVs in *SPDL1* and *KIF15*.

**Figure 5.**
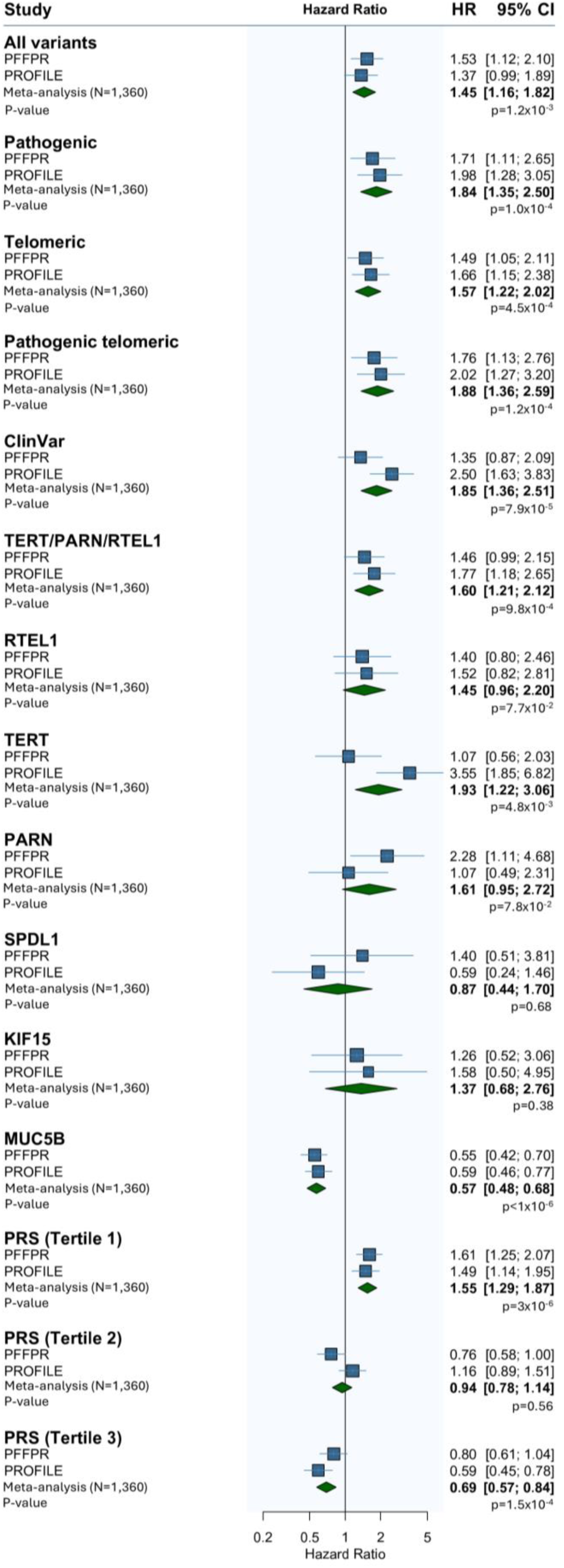
Meta-analysed results from adjusted PFFPR and PROFILE (N=1360) Cox regression models.

## DISCUSSION

Our study demonstrates that IPF patients carrying QVs in monogenic adult-onset PF genes are at increased risk of reduced survival compared to non-carriers. Additionally, we found that QV carriers tend to exhibit a lower polygenic risk component for IPF, as measured by PRS, suggesting non-additive effects between rare and common genetic variants. This indicates the existence of a distinct genetic subtype of IPF patients, defined by the interplay between these rare and common variants. These key findings were replicated across two independent studies encompassing a total of 1,360 IPF patients.

We describe for the first time that QVs in monogenic adult-onset PF genes are present in 1 in 6 to 1 in 7 IPF patients, a prevalence consistent with a recent WGS study, despite using different criteria for defining QVs (7). In comparison, our study used less stringent classification criteria for QV carrier, relying on one pathogenicity predictor instead of three, and including *SPDL1* gene, recently identified as an IPF susceptibility gene (4). Previous studies primarily focused on telomere-related genes such as *TERT, PARN, TERC*, and *RTEL1,* resulting in lower diagnostic yield (8.5-11.5% carriers of QVs) (18,30). Some earlier studies were enriched for familial IPF patients (31), who tend to carry more QVs than sporadic cases (21.4% carriers of QVs). Unlike these studies, we expanded our analysis to include additional well-defined telomere genes in the analysis, such as *DKC1, NAF1, ZCCHC8*, and *TINF2*.

Most QVs were found in telomere-related genes (75.6% in PFFPR, 68.9% in PROFILE). The association between variants in these genes and worse IPF outcomes is well-established. Carriers of QVs in *TERT, PARN, TERC*, or *RTEL1* tend to have earlier disease onset, more rapid lung function decline, and poorer survival compared to non-carriers (7,15,18), findings that are mirrored in individuals with short TL (7,17). However, the exact correlation between rare telomere-related variants and TL remains unclear, and the effect sizes of known common variants on TL are too small to fully explain this relationship (7).

Our study stands out by assessing the aggregate effect of QVs across both telomere and non-telomere genes on IPF survival. While the effect sizes were smaller compared to models that included only telomere-related variants, the most robust associations were found when considering all QVs across telomere and non-telomere genes. As expected, we identified few QVs in surfactant metabolism genes, and observed a comparable high burden of QVs in *KIF15* and *SPDL1*, two recently reported IPF genes in European cohorts. A common *SPDL1* missense variant (rs116483731) has been linked to IPF in the PROFILE cohort, particularly in the Finnish population (4), though without significant clinical differences between carriers and non-carriers. In contrast, *KIF15* rs138043992 carriers from the FinnGen FinnIPF study demonstrated early disease onset and progression (23). In the PRRPF cohort, these two risk variants were excluded from the analysis based on the applied a global genome AF cut-off. We found that carriers of *KIF15* and *SPDL1* QVs did not exhibit worse survival. However, the same direction of effect was observed among *KIF15* QV carriers in both the PFFPR and PROFILE cohorts. Future studies with larger sample sizes may be required to better define the clinical course of IPF patients with *KIF15* QVs.

Identifying prognostic markers in PF is crucial to improve the clinical management. As antifibrotic drugs like nintedanib and pirfenidone can only slow disease progression (32–34), early identification of patients with poorer prognosis could guide decisions on more aggressive treatments, such as lung transplantation. This information could also enhance the efficiency of clinical trials (35). Previous studies support that a family history of PF associates with patient survival, highlighting the importance of systematically ascertaining family histories for ILD patients to better inform prognosis (29). However, familial PF could encompass a broad group of ILD patients whose disease progression is dependent on different factors. For instance, one will be considering the risk linked to rare telomere-related variants (36,37), to common genetic variants (38), and to short age-adjusted TL (39), since they are interrelated with the family history of PF. Our findings revealed non-additive opposite effects of QVs and PRS-IPF on survival, while no apparent association was found with PRS-TL. Moreover, we did not find association between family history and IPF survival, although these results should be interpreted with caution considering that self-reported family history is prone to recall bias.

In contrast, our results show that QVs are robust predictors of IPF patient prognosis. Despite this, the use of genetic testing in IPF patients is not yet a generalized practice and current guidelines only recommend its use in familial forms of PF (9,40,41). Nonetheless we and others show the existence of a significant burden of QVs in cases in which familial history is not confirmed. It is therefore an urgent necessity to define additional criteria to improve the diagnostic yield of genetic tests in these patients. For some complex diseases, PRS have been suggested as a promising strategy for prioritizing patients who should undergo genetic sequencing (42). For example, in prostate cancer, the high penetrant variant *HOXB13* G84E is most common in cases with the lowest PRS (43). The success of this approach depends on the strength of the PRS and the genetic architecture of the disease (42). IPF meets several criteria that make it suitable for it since: 1) There is modest genetic heterogeneity in this disease and the number of susceptibility genes appears to be markedly less than other complex diseases (26); 2) Its etiology is driven by both rare, highly penetrant variants which explain monogenic-like presentations, and common variants with low effect sizes that contribute for a polygenic disorder (8); and 3) the PRS-IPF accurately identifies individuals at high risk of suffering interstitial lung abnormalities (ILA) and IPF (44). In agreement, we have found that PRS-IPF values are inversely associated with the likelihood that QVs are present in the patient. This supports the idea that PRS could serve as a valuable tool for prioritizing those patients who should undergo a deeper sequence-based analysis.

We acknowledge some limitations. First, we are aware that other genes not considered for the study are also involved in IPF risk. To test the hypothesis, the analyses were restricted to a very limited number of genes showing dominant inheritance and presumably high penetrance. This resulted in the exclusion of well-defined monogenic PF genes, such as *ABCA3* which shows recessive inheritance. In addition, there is not co-segregation data, TL measures, or functional evidence to accurately classify most QVs as P or LP. Therefore, we recognize that some variants categorized as QVs may be VUS or benign. To address this limitation, we have provided alternative QV definitions based on different *in silico* predictors and AF cutoffs which consistently showed the same direction of effect in the survival analysis. For PRS analyses, we relied on simple models based on sentinel variants from existing GWAS. For IPF, this implies that the PRS-IPF results are mainly driven by the effect of *MUC5B*. However, a recent study proved that a common genetic variant score complements the *MUC5B* variant in accurately identifying individuals at high risk of suffering ILA and IPF (44). Therefore, the use of whole-genome PRS instead of the sentinel variant-based PRS model might have benefits by offering robust associations across cohorts. Moreover, it is important to highlight that most IPF GWAS conducted to date mainly involve participants of European genetic ancestry. As a result, the PRS-IPF derived in this study might be much less accurate for non-European ancestry participants of the cohorts. Finally, despite the main findings were consistent across PFFPR and PROFILE cohorts, we acknowledge that there were differences in the definition of the primary endpoint or the received antifibrotic treatment between the two studies which could have influenced the results. Additionally, patient recruitment in PROFILE started before the approval of pirfenidone and nintedanib, which might explain the reduced median survival of patients compared to PFFPR. These differences might explain some of the inconsistencies, such as the association with survival found for *PARN* or *TERT* QV carriers only in one of the cohorts but not in the other.

## CONCLUSIONS

We found a robust association of telomere and non-telomere gene QVs with IPF patient survival. We also show that those QVs were non-additive to common IPF risk variants on survival. This study highlights the potential significance of identifying QVs in telomere and non-telomere genes linked to monogenic forms of PF in clinical practice.

## ACKNOWLEDGEMENTS

We thank all patients who participated in the PFF Patient Registry. We also thank investigators and other staff at participating PFF Care Centers for providing clinical data and blood samples, the PFF which established and has maintained the Patient Registry since 2016, and lastly, the many generous donors.

## DATA AVAILABILITY

Data supporting the findings are available as part of the manuscript or from the supplementary files. Access to the raw whole-genome sequence dataset is restricted to qualified researchers under an agreement with the PFFPR and PROFILE steering committees to protect the privacy of the participants. For further information and to apply for access to the data from the PFFPR prior to public deposit in BioLINCC, please contact the chair of the ancillary committee (Dr. Noth) or any other member of the steering committee (https://www.pulmonaryfibrosis.org/pff-registry/pff-patient-registry). For further information and to apply for access to the PROFILE data, please contact. Data access requests must be reviewed before release.

## AUTHOR CONTRIBUTIONS

Conceptualization and supervision: A.A.G., I.N., and C.F. Patient recruitment, collection of biospecimen, and clinical data: J.M.O, P.L.M., J.A.K., A.A., F.J.M., T.M.M., T.S.B., R.G.J., I.N., I.S., B.Y., S.F.M., E.S., J.M., J.S.K., Y.H., L.V.W., R.J.A., D.Z., and C.G. Formal analysis: A.A.G., D.J., J.M.L.S., and D.Z. Supervision: I.N. and C.F. Writing-original draft: A.A.G. and C.F. Funding acquisition: C.F., L.V.W., and I.N. Visualization: A.A.G. Writing-review and editing: All the authors.

## Supplementary material

## Supplementary methods

### Description of study cohorts

The Pulmonary Fibrosis Foundation Patient Registry (PFFPR) is a large multicentre based registry that collects baseline and longitudinal demographic and clinical information about well-characterized patients with interstitial lung diseases (ILD) in the United States since March 2016 to allow retrospective and prospective research^1^. In addition, the PFFPR major objective is to apply blood-based omics technologies (whole-genome sequencing [WGS], proteomic analysis, and transcriptional profiling) on blood samples from patients to study molecular markers of the onset or progression of diseases. Patients aged ≥18 years old who has ILD diagnosed and had not undergone lung transplantation were recruited from approximately 42 USA sites selected primarily from the familial pulmonary fibrosis (FPF) Care Center Network.

They were followed for the progression of the disease through the lifetime of the PFFPR or the patient until the patient receives lung transplant. More details of the PFFPR including inclusion and exclusion criteria as well as collected clinical variables are described elsewhere^1^. The PFFPR cohort includes 1317 individuals with ILD for whom WGS data are available. For this study, we included the 917 PFFPR patients with a definitive IPF diagnosis. Family history was available for all of them although no genetic causes were previously assessed. After the quality control procedures, 888 of those patients remained in the study **(Figure 1)**.

The PROFILE is a UK large, prospective, multicentre, longitudinal study conducted on patients with fibrotic ILD^2,3^. The cohort includes 541 patients with IPF or idiopathic non-specific interstitial pneumonia aged 18-85 recruited from tertiary specialist ILD and from local secondary care hospitals. Blood samples for genomic analysis were collected and they were followed for disease progression through 3 years. After quality control steps, the second stage of the study included 472 patients with a confirmed diagnosis of IPF **(Figure 1).**

Baseline characteristics of the PFFPR and the PROFILE cohorts are listed in **Supplementary Table 1**.

### Supplementary bioinformatics methods

In both cohorts, several quality control (QC) analyses were performed: (i) detection of QC outliers, (ii) the kinship between patients, (iii) sample cross-contamination, and (iv) sex discordance. We used a combination of DRAGEN metrics, and assessments with PLINK v1.90b6.24^4^, SCE-VCF v0.1.2 (https://github.com/HTGenomeAnalysisUnit/SCE-VCF), Somalier v0.2.19^5^, and KING v2.3.2^6^.

#### Detection of QC outliers

Based on PLINK analysis, we detected abnormal heterozygosity rate and genotyping call rate to infer potential sample contaminations and/or a low DNA concentration. A heterozygosity rate value ± 3 standard deviations from the mean and/or genotype call rate <0.95 were considered as outliers.

#### Kinship between patients

We detected duplicates or monozygotic twins, and first-degree kinship relationships with three different tools: we considered two samples as duplicates or obtained from monozygotic twins if a PI_HAT value was >0.9 for PLINK, a Somalier relatedness value >0.9, and a KING kinship coefficient >0.354. We considered as first-degree relatives a PI_HAT in the range of 0.4-0.6 for PLINK, a Somalier relatedness value in the range of 0.4-0.6, and a KING kinship coefficient in the range of 0.177-0.354. We found a complete consensus among these tools in the cohort. Second-degree relatives were not detected.

#### Sample cross-contamination

We used the “*estimated_sample_contamination*” parameter from DRAGEN metrics to exclude samples with evidence ≥2% of contamination. We also used SCE-VCF tool, which estimates contamination from VCF files using the CHARR method^7^, based on the recommended thresholds to consider a sample as contaminated (CHARR > 0.03 and INCONSISTENT_AB_HET_RATE > 0.15). We found a complete consensus among these tools in the identification of potential sample contamination in the PFF-PR. For PROFILE, we only relied on SCE-VCF for the sample cross-contamination inference.

#### Sex discordance

Biological sex inference from genetic data was obtained with Somalier following recommendations. For that we compared the scaled mean depth on X and Y chromosomes for 365 and 17 genomic positions, respectively. Sex discordance, identified by comparing the genetically inferred sex with that recorded, was also used to exclude patients from the study. In the PFF-PR, a female was identified as a possible X0 aneuploid due to the low number of heterozygous sites on the X chromosome and was excluded from the analysis.

## Supplementary results

### Prevalence of qualifying variants (QV) in PROFILE

The genes with the highest burden of QVs were: *RTEL1* (20.5%), *TERT* (15.1%), and *PARN* (17.8%) **(Supplementary Figure 2B**, **2D)**. The prevalence of QVs among carriers of the risk *MUC5B* genotype (rs35705950-T) was lower (14.97%) than among those carrying the protective GG genotype (16.85%), although the difference was not statistically significant (*p*=0.60). We observed the same effect direction as in PFFPR when assessing the association between the lower PRS-IPF tertile and reduced survival (HR=1.49, 95% CI=0.14-1.95, *p*=3.1×10^-3^) **(Figure 3B)**.

## Supplementary tables

**Supplementary Table 1.**
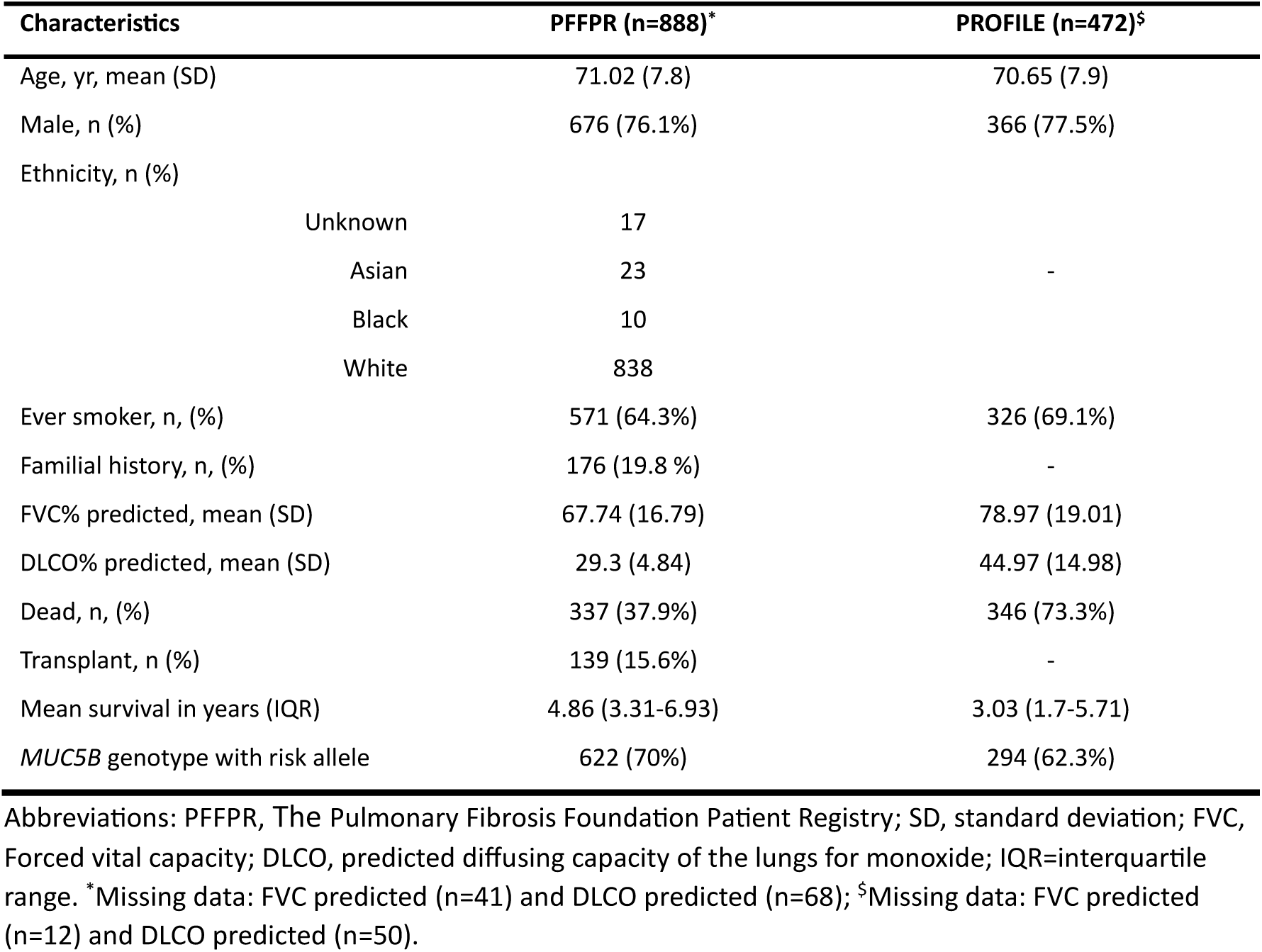
Baseline characteristics and outcomes of IPF patients from stage one and stage two cohorts.

| Characteristics | PFFPR (n=888)* | PROFILE (n=472)<sup>§</sup> |
| --- | --- | --- |
| Age, yr, mean (SD) | 71.02 (7.8) | 70.65 (7.9) |
| Male, n (%) | 676 (76.1%) | 366 (77.5%) |
| Ethnicity, n (%) |  |  |
| Unknown | 17 |  |
| Asian | 23 | - |
| Black | 10 |  |
| White | 838 |  |
| Ever smoker, n, (%) | 571 (64.3%) | 326 (69.1%) |
| Familial history, n, (%) | 176 (19.8 %) | - |
| FVC% predicted, mean (SD) | 67.74 (16.79) | 78.97 (19.01) |
| DLCO% predicted, mean (SD) | 29.3 (4.84) | 44.97 (14.98) |
| Dead, n, (%) | 337 (37.9%) | 346 (73.3%) |
| Transplant, n (%) | 139 (15.6%) | - |
| Mean survival in years (IQR) | 4.86 (3.31-6.93) | 3.03 (1.7-5.71) |
| *MUC5B* genotype with risk allele | 622 (70%) | 294 (62.3%) |

Abbreviations: PFFPR, The Pulmonary Fibrosis Foundation Patient Registry; SD, standard deviation; FVC, Forced vital capacity; DLCO, predicted diffusing capacity of the lungs for monoxide; IQR=interquartile range. \*Missing data: FVC predicted (n=41) and DLCO predicted (n=68); <sup>§</sup>Missing data: FVC predicted (n=12) and DLCO predicted (n=50).

**Supplementary Table 2.** Regions of interest (ROIs) for qualifying variants annotations in hg38.

| Chromosome | Gene | Start | End |
| --- | --- | --- | --- |
| 5 | *TERT* | 1,253,047 | 1,295,168 |
| 3 | *TERC* | 169,764,420 | 169,765,160 |
| 14 | *TINF2* | 24,238,186 | 24,242,763 |
| X | *DKC1* | 154,762,642 | 154,777,789 |
| 20 | *RTEL1* | 63,657,710 | 63,696,353 |
| 16 | *PARN* | 14,435,600 | 14,632,828 |
| 4 | *NAF1* | 163,109,973 | 163,166,990 |
| 12 | *ZCCHC8* | 122,471,500 | 122,501,032 |
| 8 | *SFTPC* | 22,156,813 | 22,164,579 |
| 10 | *SFTPA2* | 79,555,752 | 79,560,507 |
| 10 | *SFTPA1* | 79,610,839 | 79,615,555 |
| 5 | *SPDL1* | 169,583,536 | 169,604,878 |
| 3 | *KIF15* | 44,761,621 | 44,873,476 |

**Supplementary Table 3.**
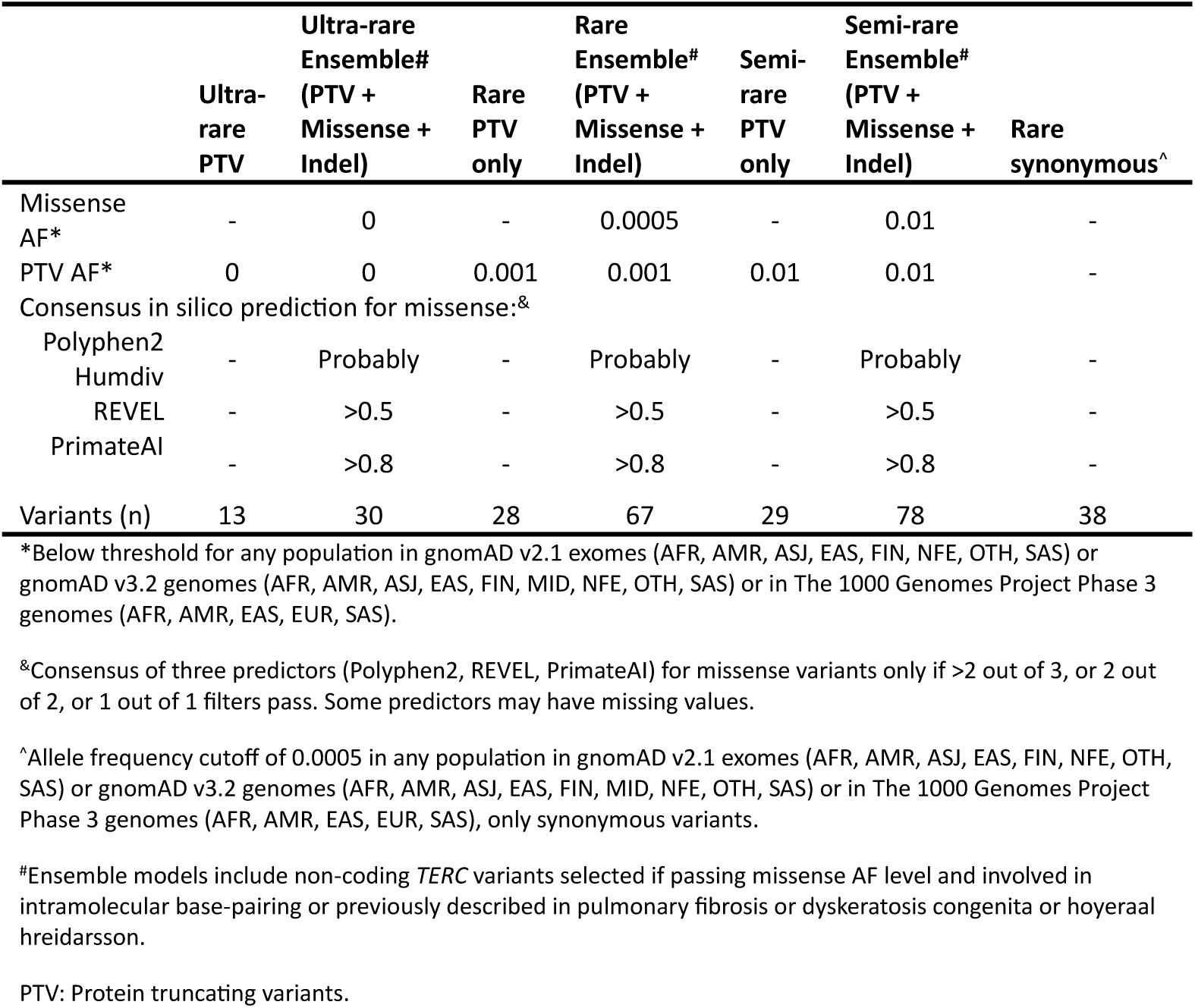
Alternative definitions for qualifying variants and the rare synonymous used for sensitivity analyses.

**Supplementary Table 4.**
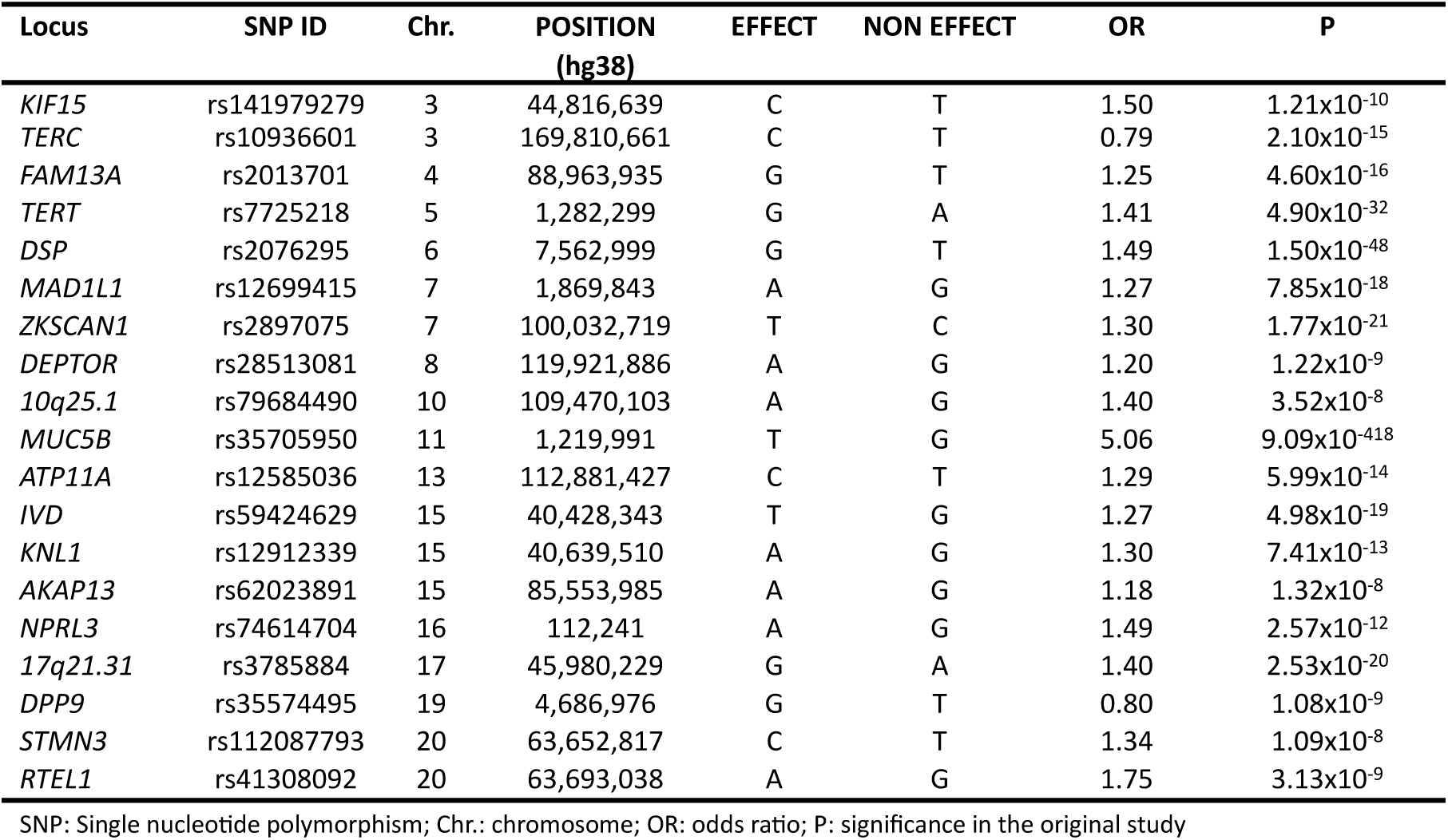
Common IPF risk variants and effects considered for PRS-IPF estimation.

**Supplementary Table 5.**
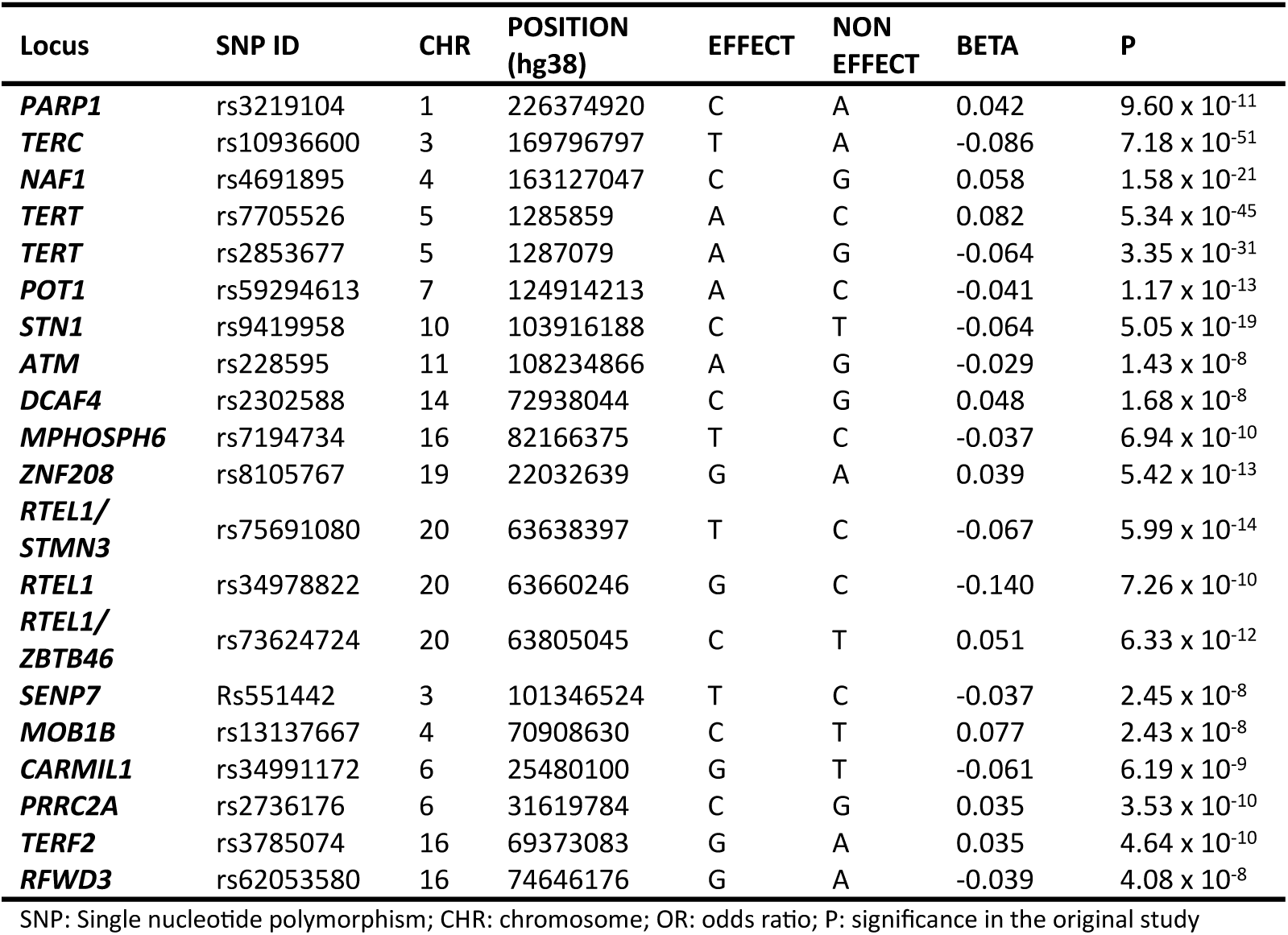
Common telomere length variants and effects considered for PRS-TL estimation.

## Supplementary Figures

**Supplementary Figure 1.**
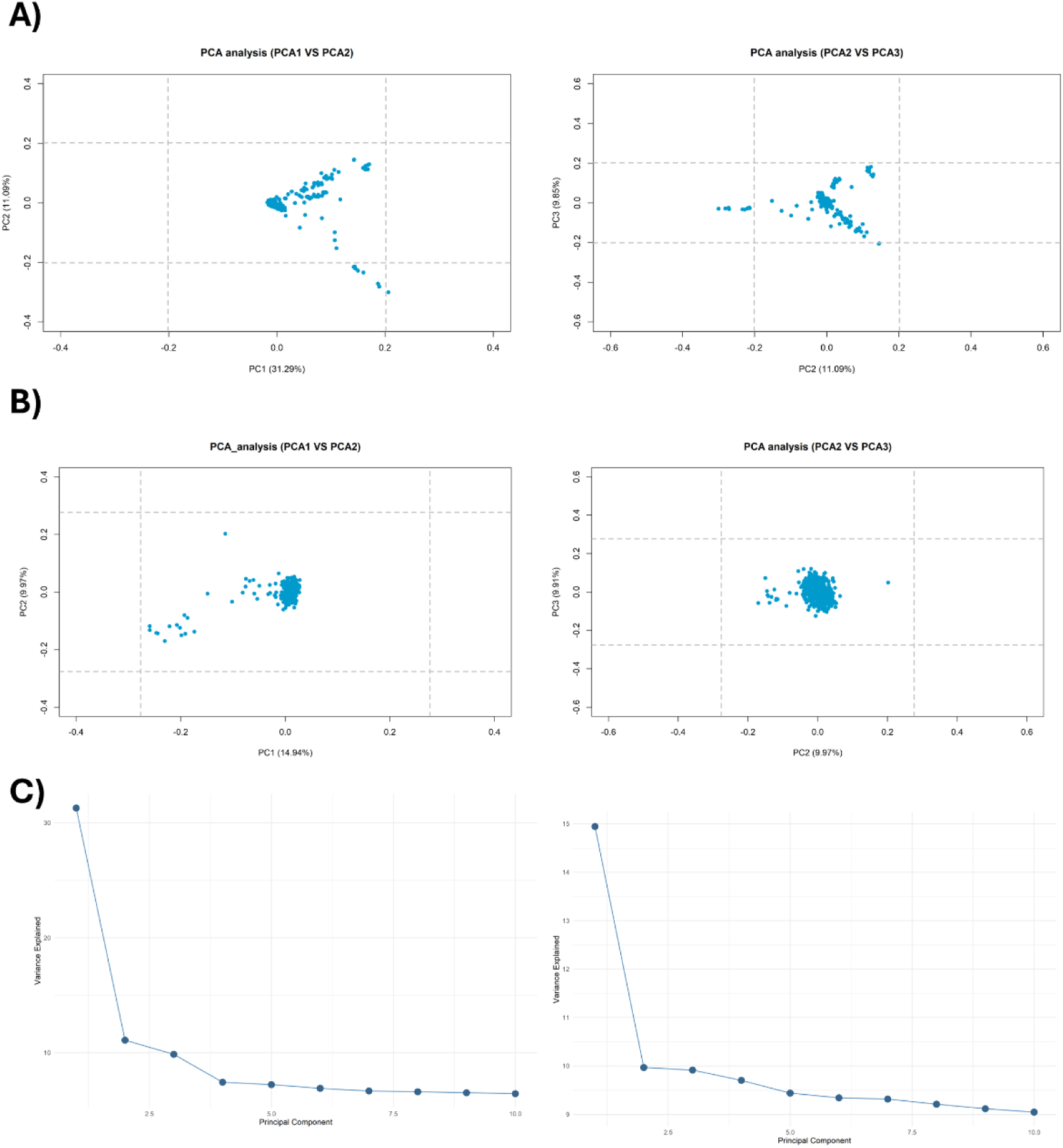
Principal component analysis. A) Plot of the first two (left) and the second and third (right) principal components of genetic variation of IPF patients in the PFFPR. B) Plot of the first two (left) and the second and third (right) principal components of genetic variation of IPF patient in PROFILE. C) Proportion of variance explained by each PC (PFFPR on the right, and PROFILE on the left).

**Supplementary Figure 2.**
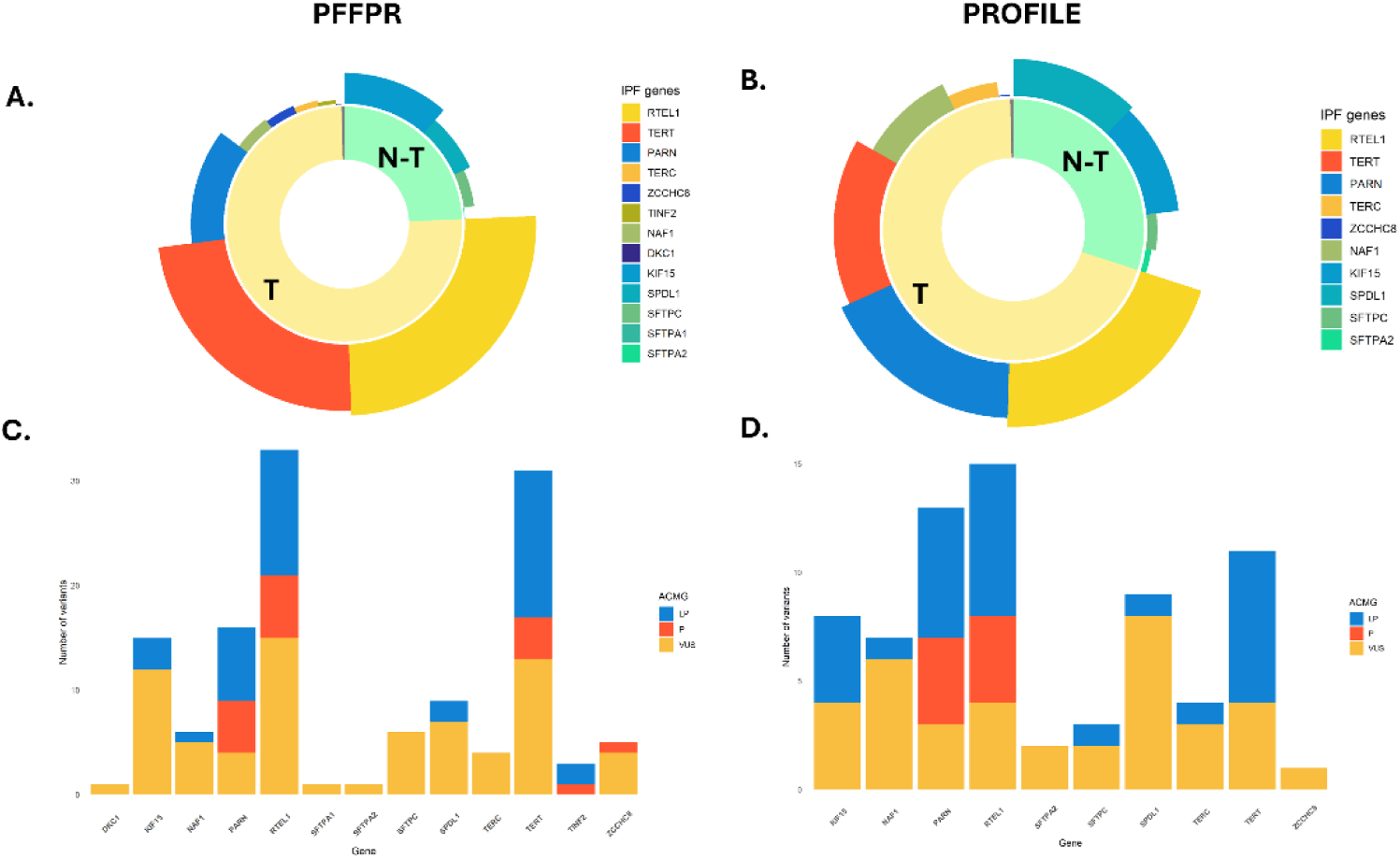
Distribution of qualifying variants (QV) in monogenic adult-onset pulmonary fibrosis (PF) genes in the PFFPF and PROFILE cohorts. A) Total QVs in monogenic adult-onset PF genes in the PFFPR. B) Total QVs in monogenic adult-onset PF genes in the PROFILE cohort. C) Variants classified in P/LP/VUS per gene in the PFFPR. D) Variants classified in P/LP/VUS per gene in the PROFILE cohort. T: Telomere; N-T: Non telomere.

**Supplementary Figure 3.**
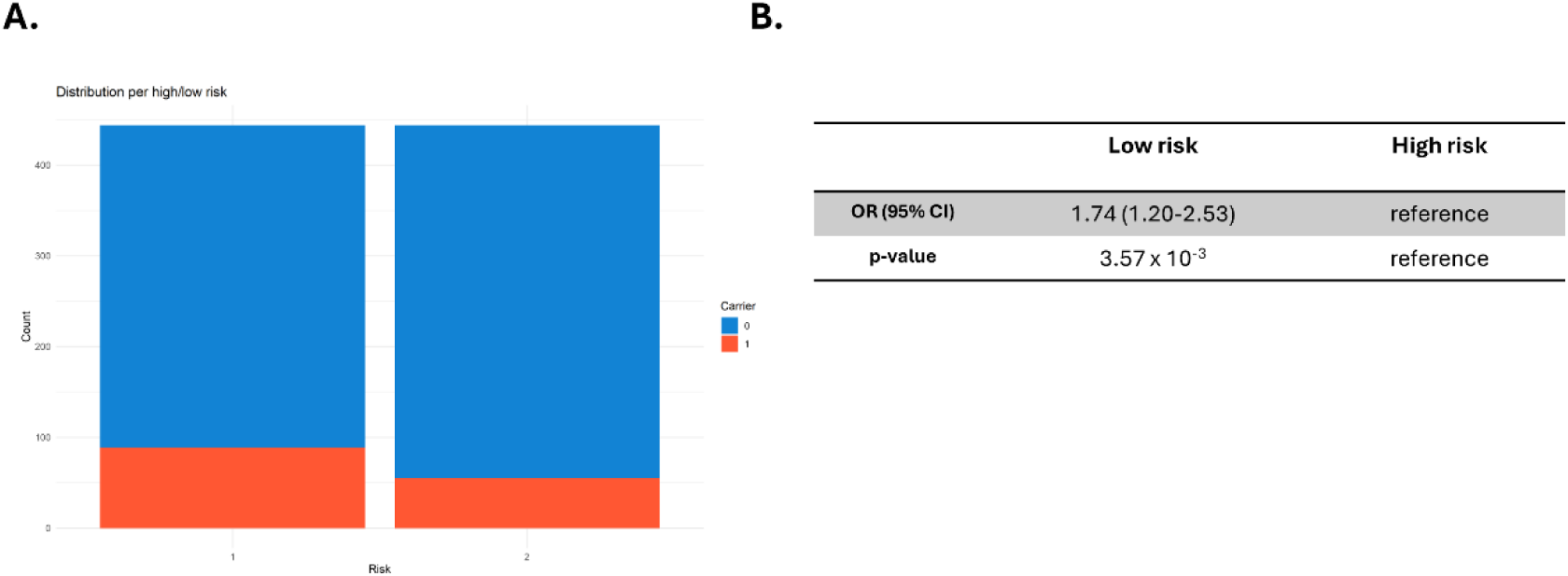
Association between prevalence of qualifying variants (QV) and PRS-IPF in the PFFPR. A) Distribution of carriers (1) and non-carriers (0) in low and high PRS-IPF. B) Risk of carrying a QV in patients with low polygenic risk in comparison with individuals with high polygenic risk. The odds ratio (OR) and the 95% confidence interval (CI) were estimated using logistic regression adjusted by age of diagnosis, sex, and the two main principal components.

**Supplementary Figure 4.**
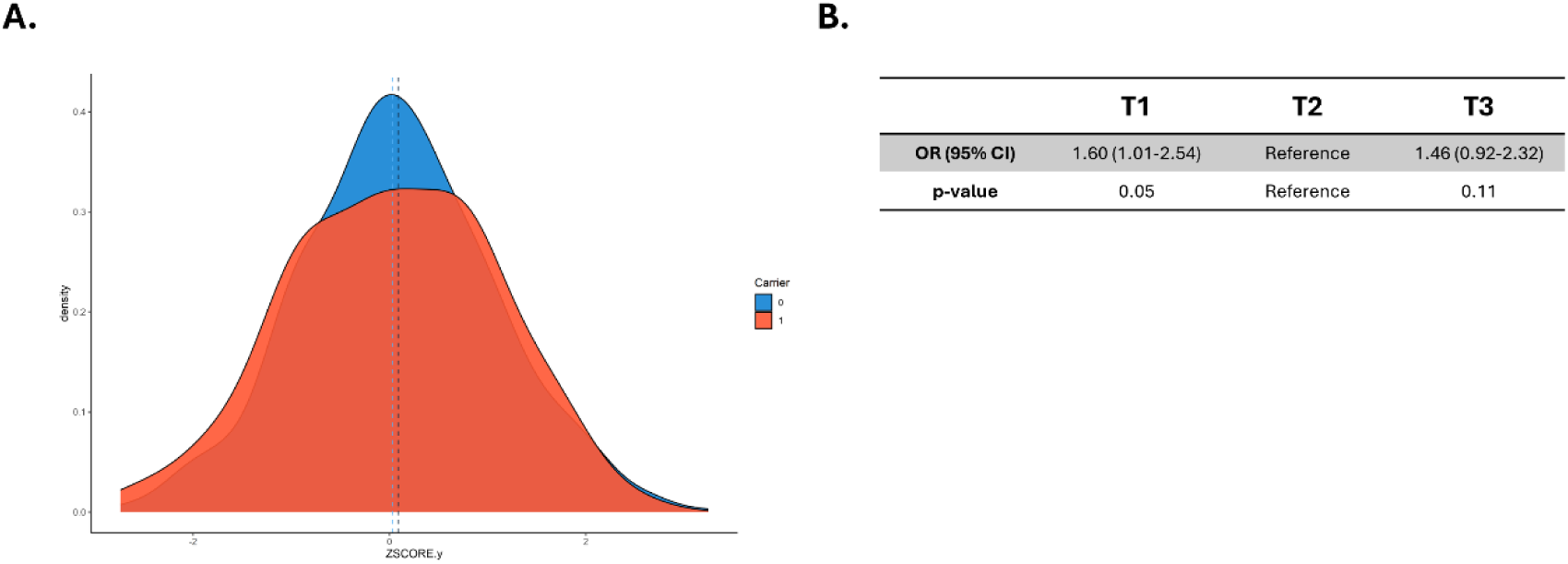
Association between prevalence of qualifying variants (QV) and PRS-IPF (after excluding the *MUC5B* locus) in the PFFPR. A) Distribution of PRS-IPF in carriers (1) and non-carriers (0). Vertical dotted lines represent the mean value of the distribution. B) Risk of carrying a QV for patients with low polygenic risk (T1) and high polygenic risk (T3) compared to those in the middle tertile. The odds ratios (OR) and the 95% confidence intervals (CI) were estimated using logistic regression adjusted for age of diagnosis, sex, and the two main principal components.

**Supplementary Figure 5.**
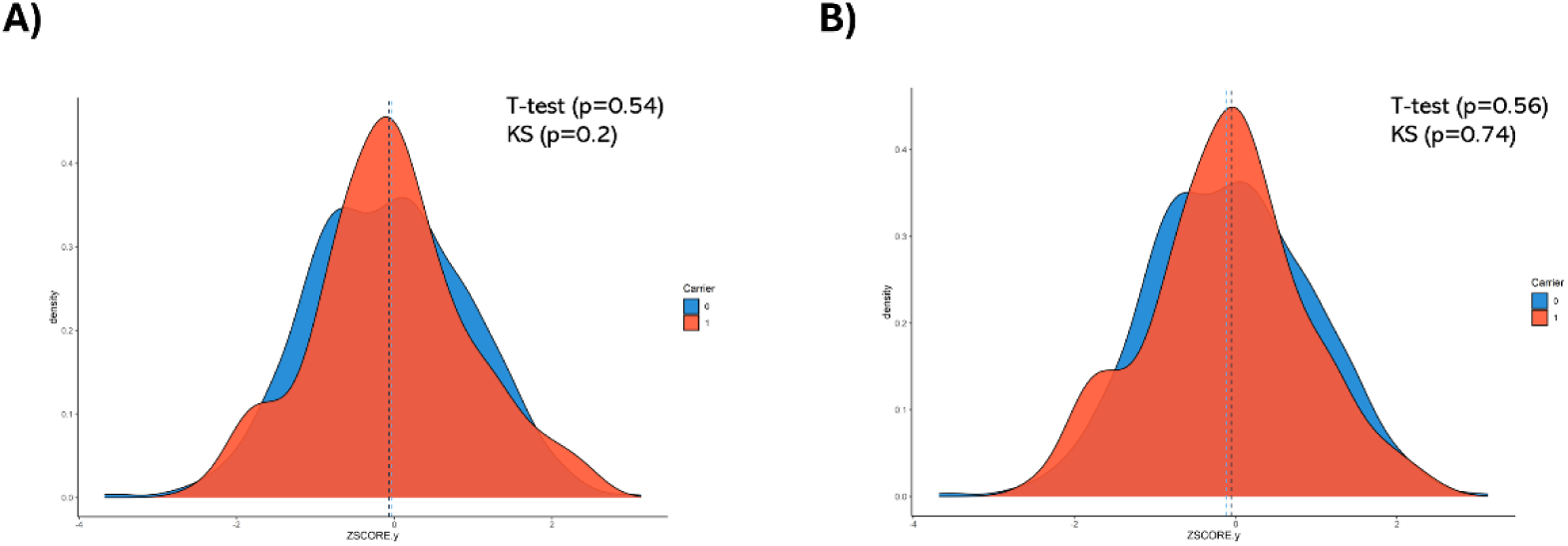
Association between the prevalence of qualifying variants (QV) and PRS-TL in the PFFPR. Distribution of PRS-TL in carriers (1) and non-carriers (0). Vertical dotted lines represent the mean value of the distribution A) Carriers (1) and non-carriers (0) in telomere and non-telomere genes. B) Carriers (1) and non-carriers (0) in telomere genes. T-test: Student’s t-test; KS: Kolmogorov-Smirnov test.

**Supplementary Figure 6.**
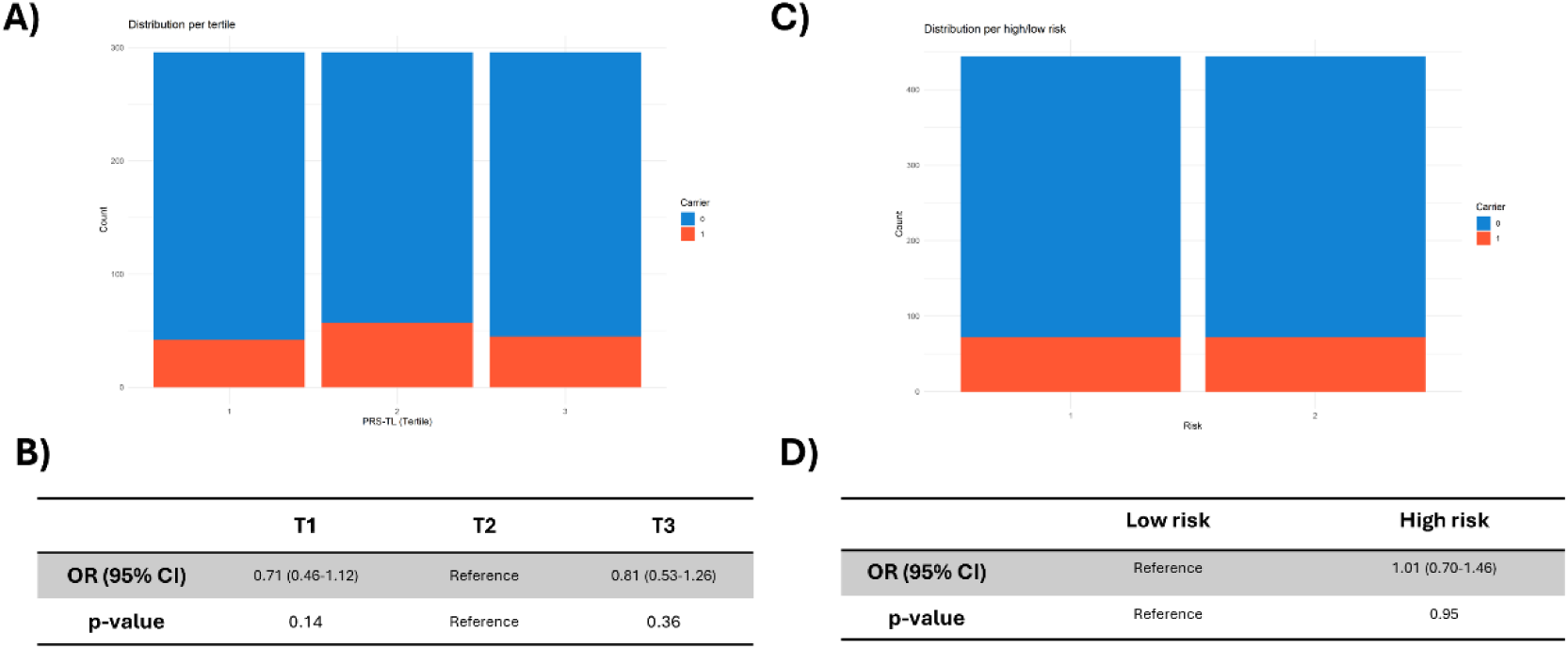
Association between prevalence of qualifying variants (QV) in telomere and non-telomere genes and PRS-TL in the PFFPR. A) Distribution of carriers (1) and non-carriers (0) in PRS-TL tertiles. B) Risk of carrying a QV for individuals with low polygenic risk (T1) and high polygenic risk (T3) compared to those in the middle tertile. C) Distribution of carriers (1) and non-carriers (0) in high and low PRS-TL. D) Risk of carrying a QV in patients with high polygenic risk in comparison with patients with low polygenic risk. The odds ratios (OR) and the 95% confidence intervals (CI) were estimated using logistic regression adjusted by age of diagnosis, sex, and the two main principal components.

**Supplementary Figure 7.**
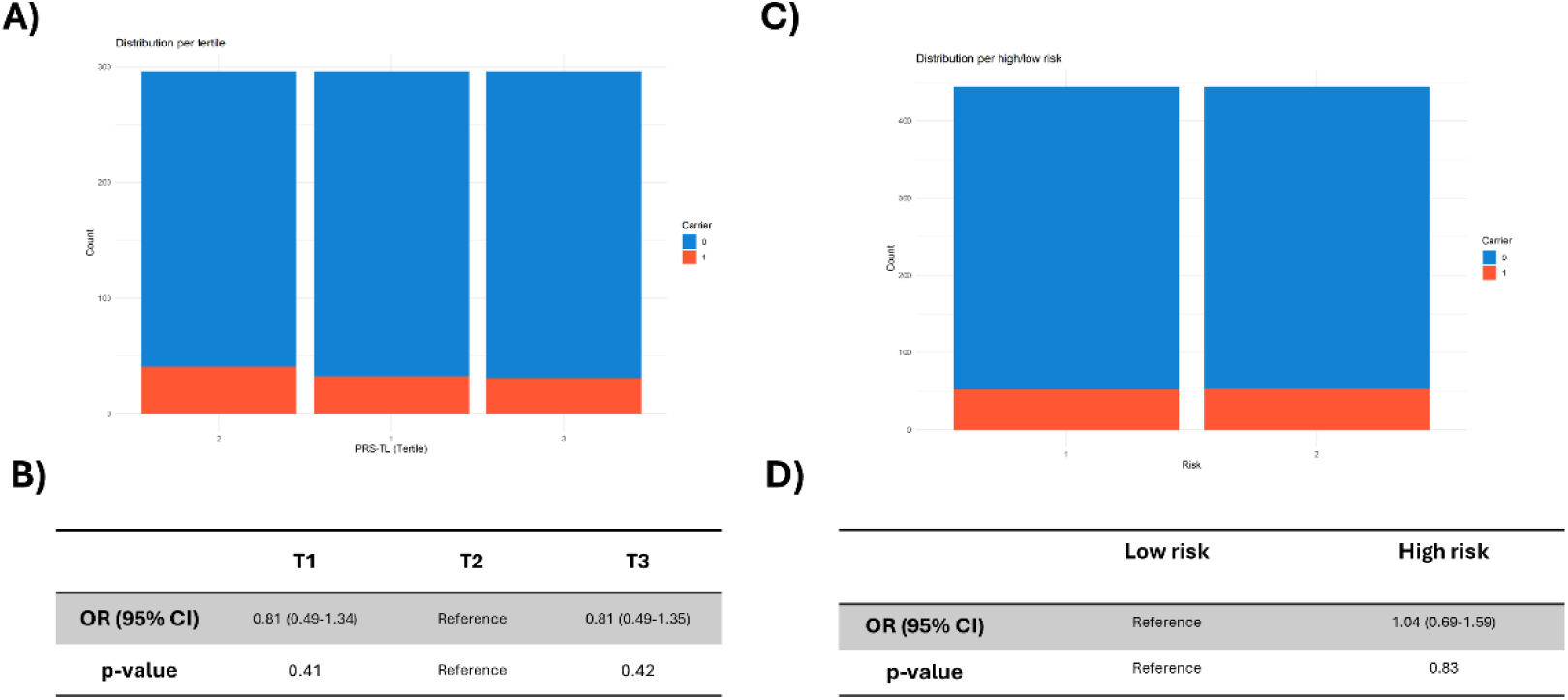
Association between prevalence of qualifying variants (QV) in telomere genes and PRS-TL in the PFFPR. A) Distribution of carriers (1) and non-carriers (0) in PRS-TL tertiles. B) Risk of carrying a QV for individuals with low polygenic risk (T1) and high polygenic risk (T3) compared to those in the middle tertile. C) Distribution of carriers (1) and non-carriers (0) in high and low PRS-TL. D) Risk of carrying a QV in individuals with high polygenic risk in comparison with individuals with low polygenic risk. The odds ratios (OR) and the 95% confidence intervals (CI) were estimated using logistic regression adjusted by age of diagnosis, sex, and the two main principal components.

**Supplementary Figure 8.**
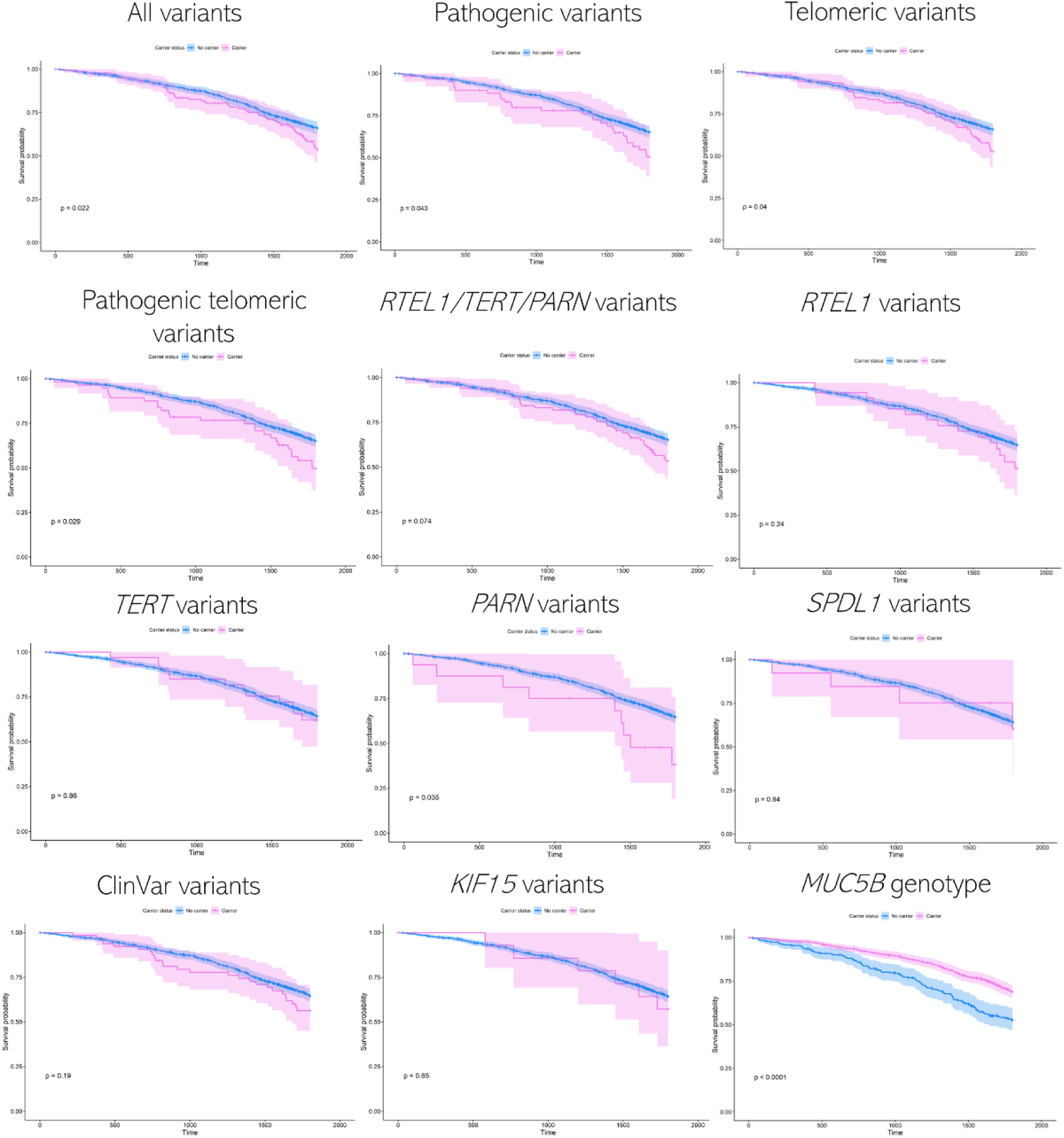
Kaplan-Meier survival analysis for qualifying variants (QV) (per gene and group of genes) and the *MUC5B* risk allele in the PFFPR. p-values for the log-rank test are shown.

**Supplementary Figure 9.**
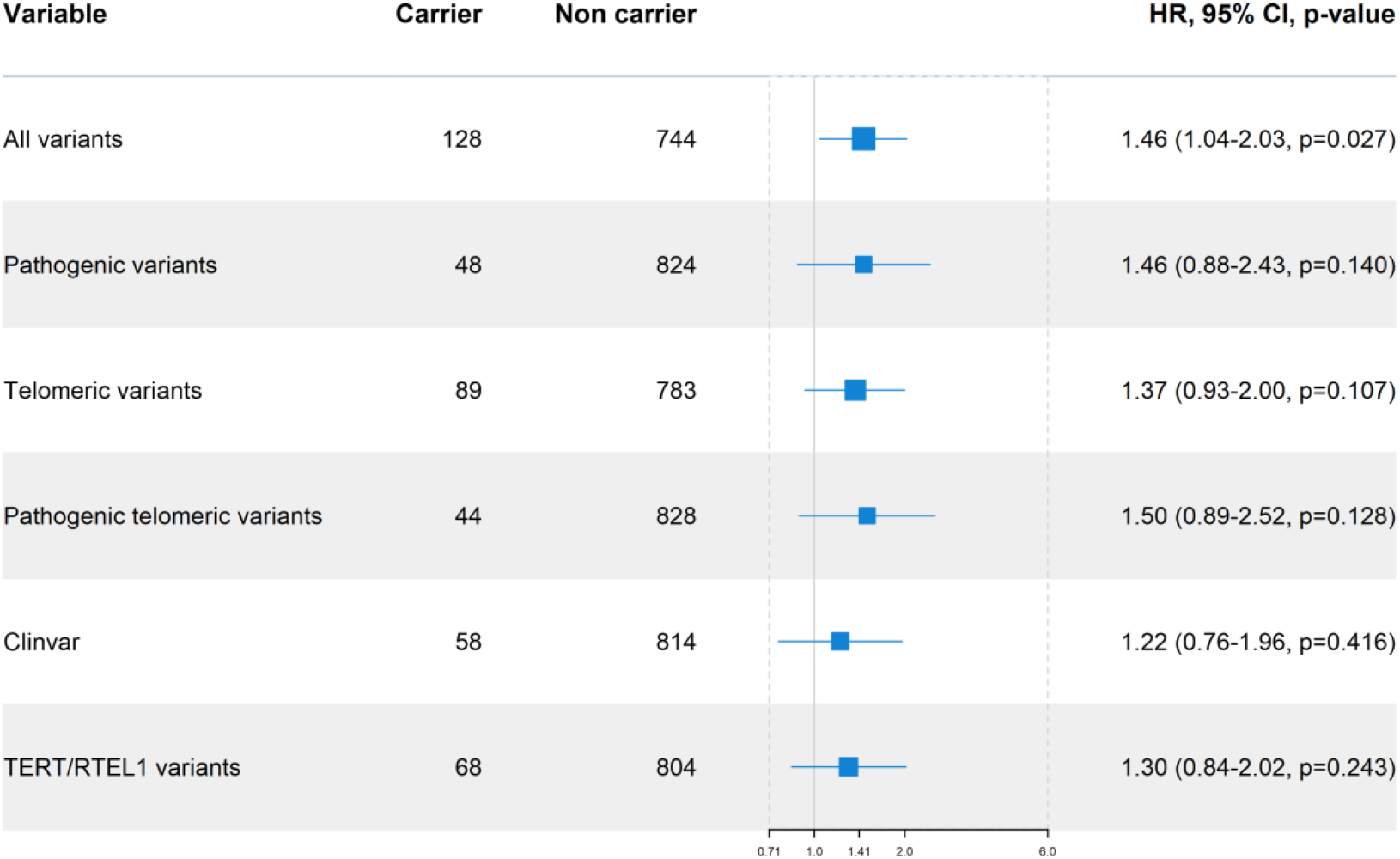
Qualifying variants (QV) effect on survival in the PFFPR (excluding carriers of QV within *PARN*). All analysis were performed using Cox regression models adjusted for sex, age of diagnosis, the two main principal components, *MUC5B* risk allele, smoking history, forced vital capacity (FVC) % predicted, and diffusing capacity for carbon monoxide (DLCO) % predicted. The X-axis shows Hazard-ratios (HR); the grey line corresponds to the HR=1.0. The boxes correspond to adjusted HR and horizontal lines correspond to 95% confidence intervals (CI).

**Supplementary Figure 10.**
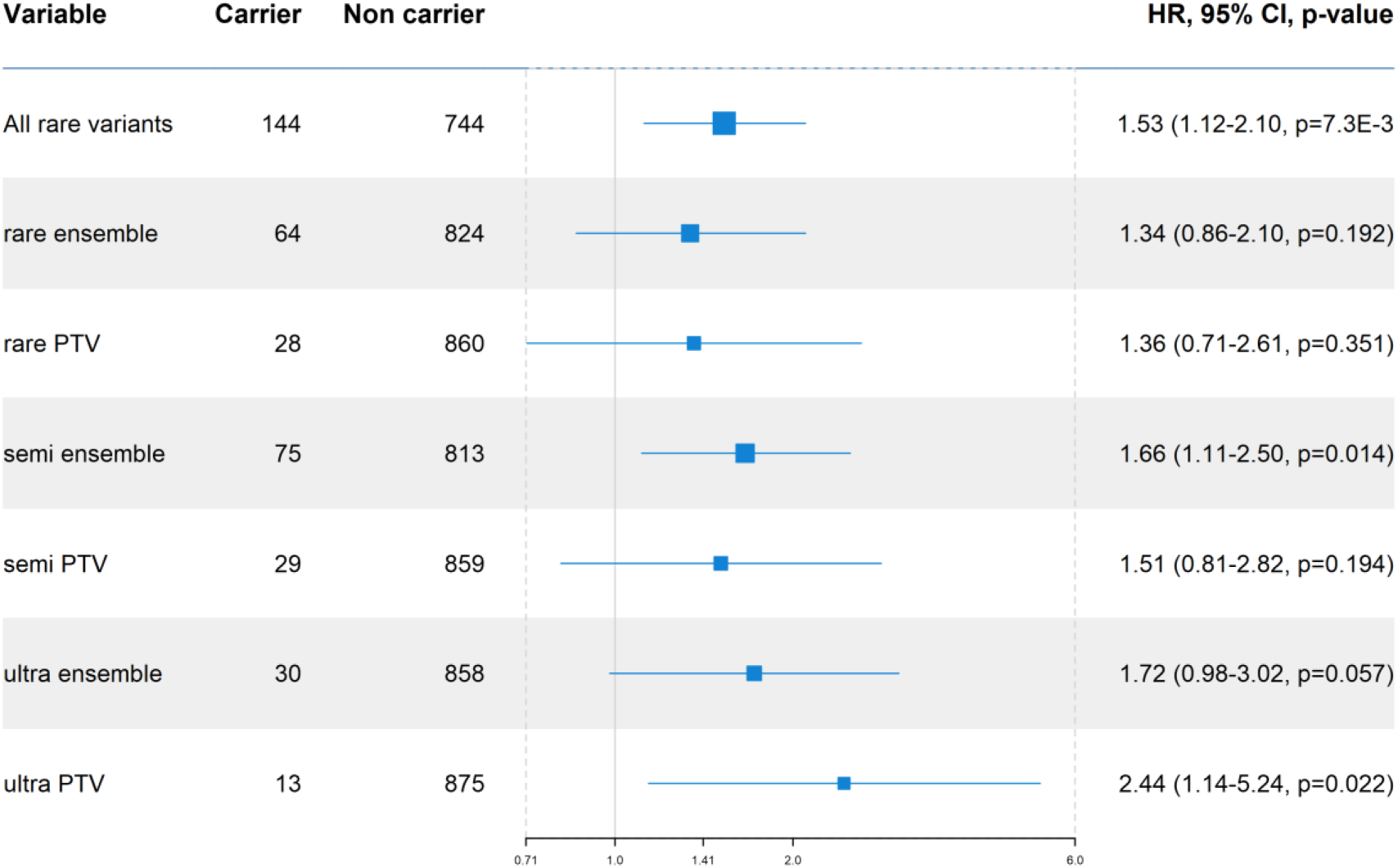
Alternative qualifying variants (QV) classifications and effects on survival in the PFFPR. All analysis were performed using Cox regression models adjusted for sex, age of diagnosis, the two main principal components, *MUC5B* risk allele, smoking history, forced vital capacity (FVC) % predicted, and diffusing capacity for carbon monoxide (DLCO) % predicted. The X-axis shows Hazard-ratios (HR); the grey line corresponds to the HR=1.0. The boxes correspond to adjusted HR and horizontal lines correspond to 95% confidence intervals (CI).

**Supplementary Figure 11.**
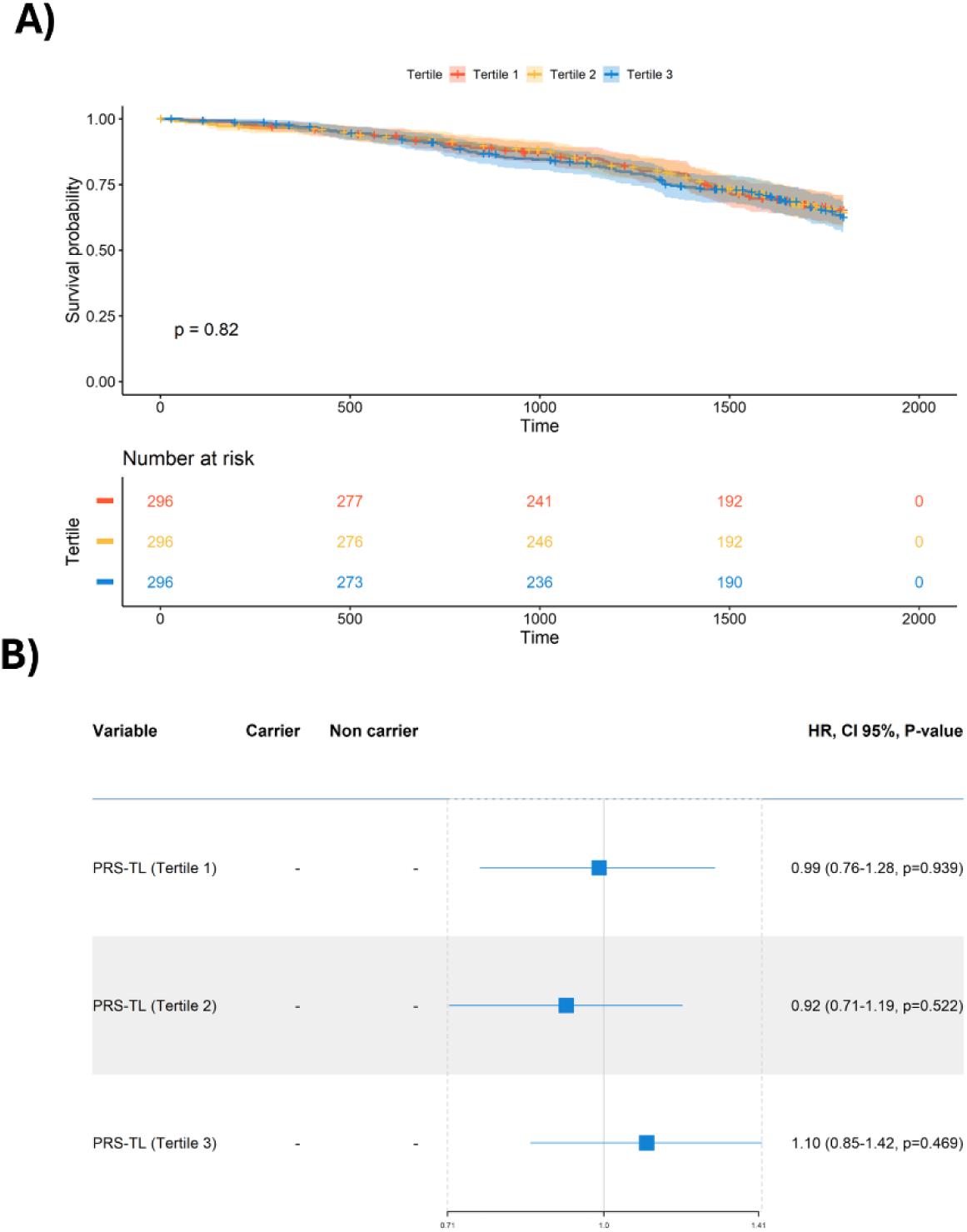
Association between PRS-TL tertiles and survival in the PFFPR. A) Kaplan-Meier survival analysis for PRS-TL tertiles (p-value for the log-rank test is shown). B) PRS-TL effect on survival. The analysis was performed using Cox regression models adjusted for sex, age of diagnosis, the two main principal components, smoking history, forced vital capacity (FVC) % predicted, and diffusing capacity for carbon monoxide (DLCO) % predicted. The X-axis shows Hazard-ratios (HR); the grey line corresponds to the HR=1.0. The boxes correspond to adjusted HR and horizontal lines correspond to 95% confidence intervals (CI).

**Supplementary Figure 12.**
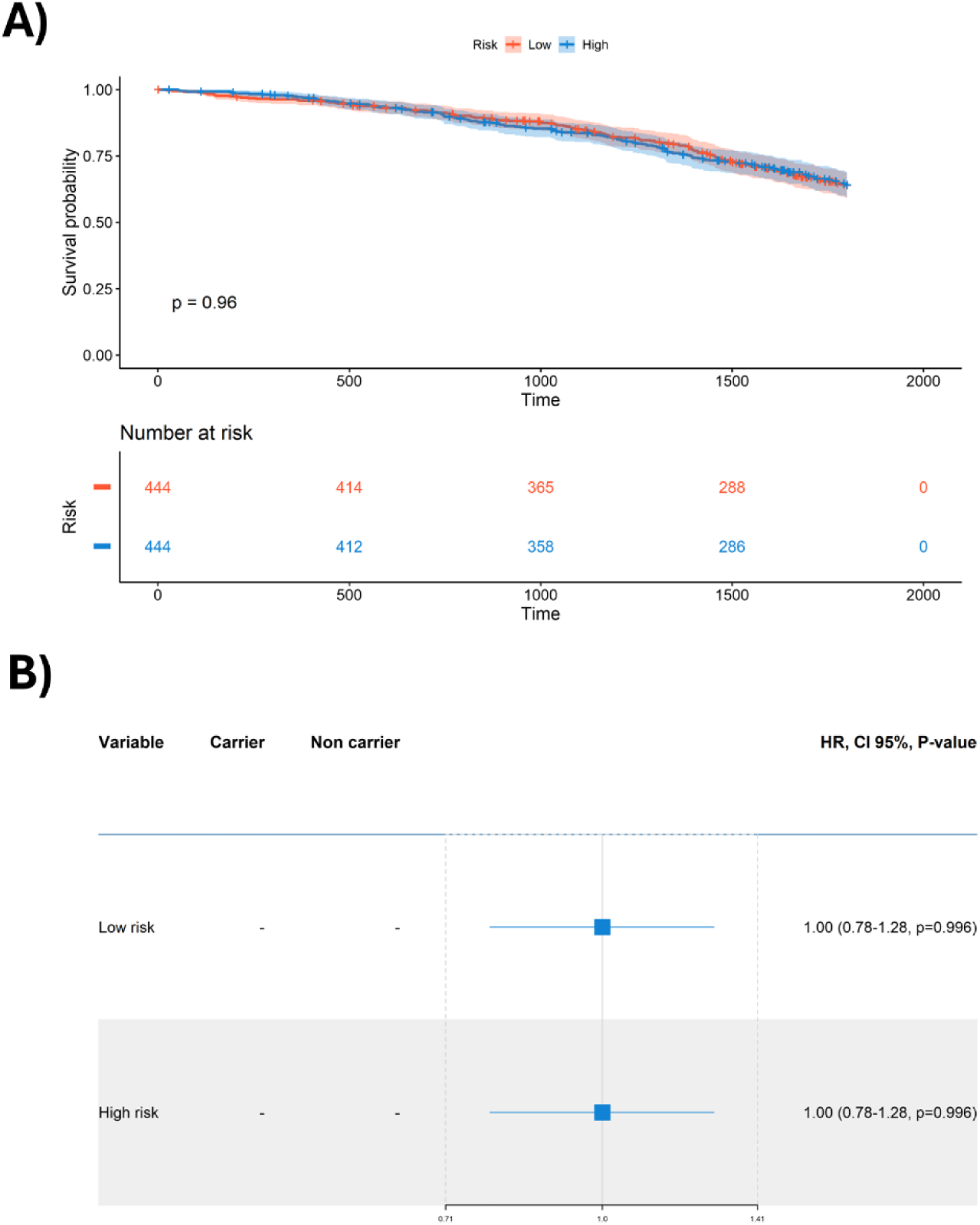
Association between high and low PRS-TL and survival in the PFFPR. A) Kaplan-Meier survival analysis for high/low risk PRS-TL (p-value for the log-rank test is shown). B) PRS-TL effect on survival. The analysis was performed using Cox regression models adjusted for sex, age of diagnosis, the two main principal components, smoking history, forced vital capacity (FVC) % predicted, and diffusing capacity for carbon monoxide (DLCO) % predicted. The X-axis shows Hazard-ratios (HR); the grey line corresponds to the HR=1.0. The boxes correspond to adjusted HR and horizontal lines correspond to 95% confidence intervals (CI).

**Supplementary Figure 13.**
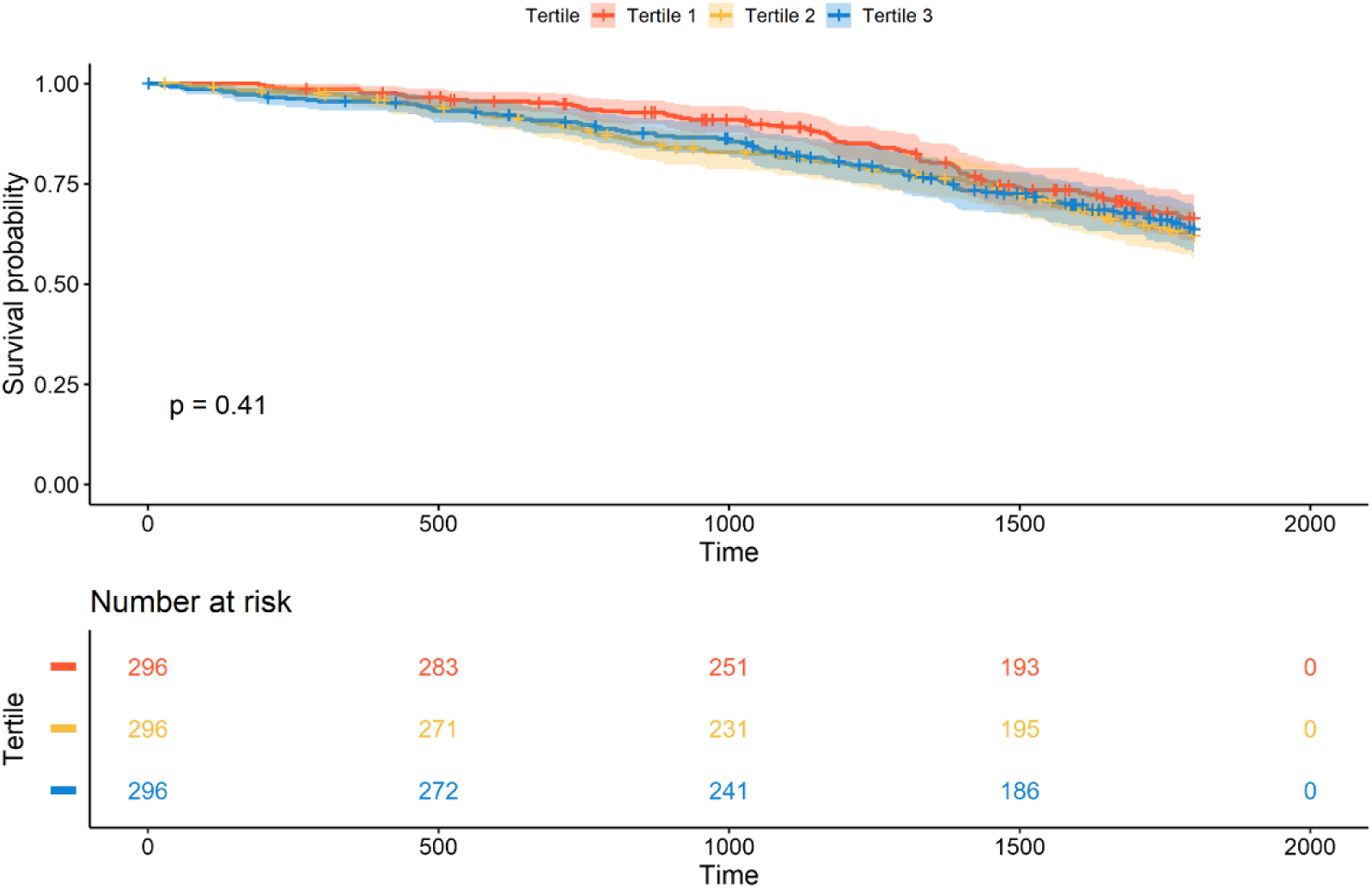
Association of PRS-IPF (after excluding the *MUC5B* locus) and survival in the PFFPR. Kaplan-Meier analysis showing p-values for the log-rank test.

**Supplementary Figure 14.**
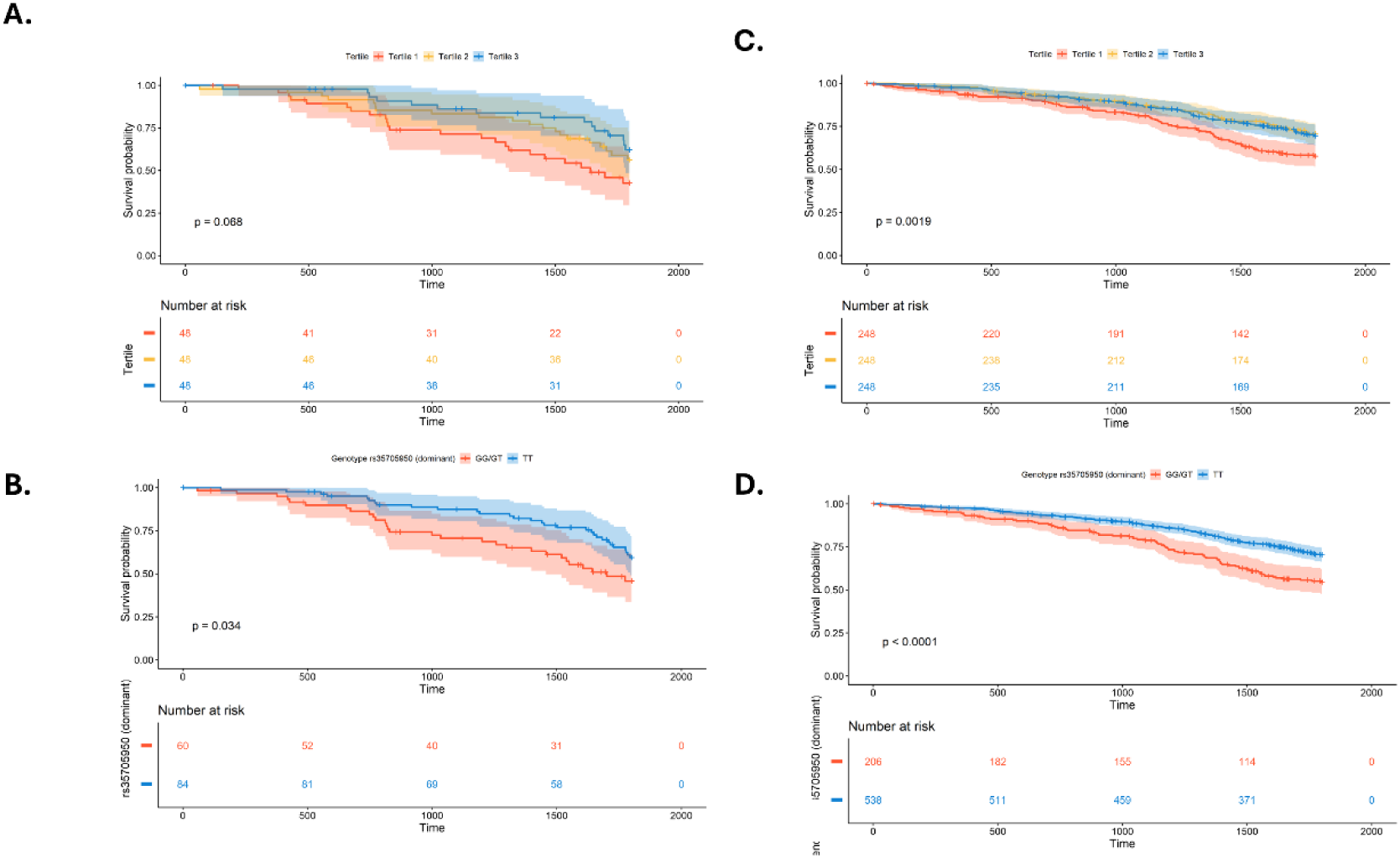
Associations between PRS-IPF and *MUC5B* rs35705950 genotypes with survival among carriers and non-carriers of qualifying variants (QV) in the PFFPR. A) Association between PRS-IPF and survival in carriers. B) Association between PRS-IPF and survival in non-carriers. C) Association between *MUC5B* rs35705950 genotypes and survival in carriers. D) Association between *MUC5B* rs35705950 genotypes and survival in non-carriers. Kaplan-Meier analysis, showing p-values for the log-rank test.

**Supplementary Figure 15.**
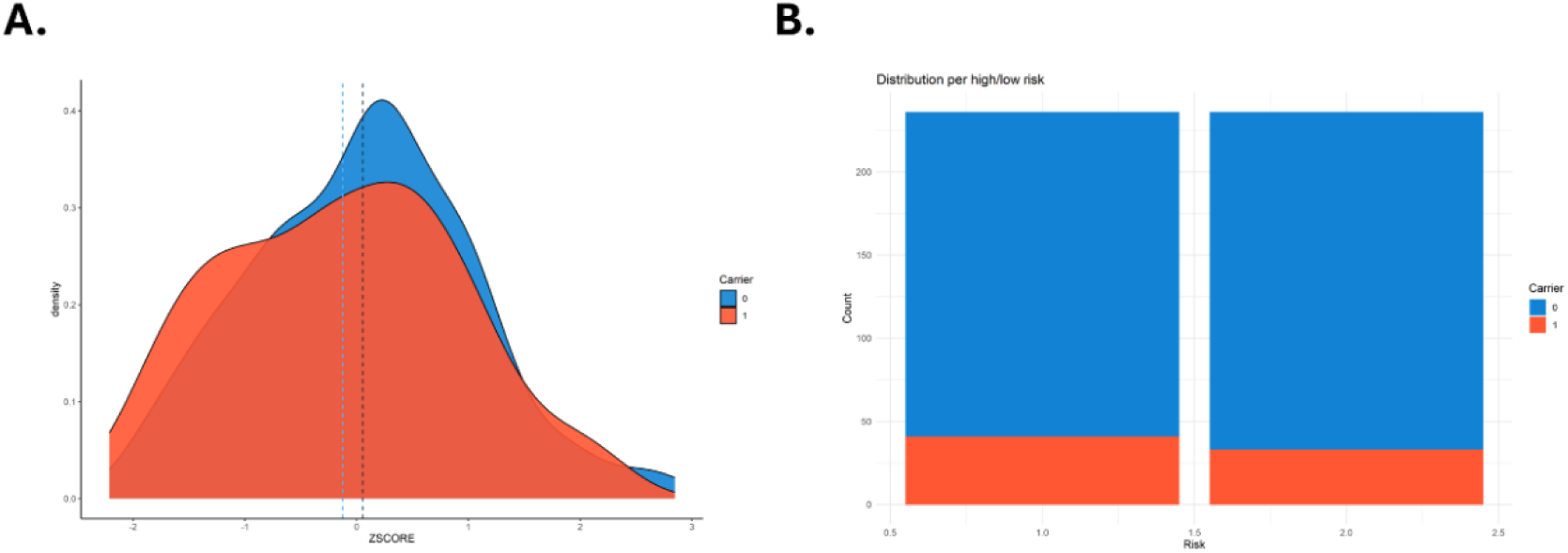
Association between prevalence of qualifying variants (QV) and PRS-IPF in PROFILE. A) Distribution of PRS-IPF in carriers (1) and non-carriers (0). Vertical dotted lines represent the mean value of the distribution. B) Distribution of carriers (1) and non-carriers (0) in high and low PRS-IPF.

**Supplementary Figure 16.**
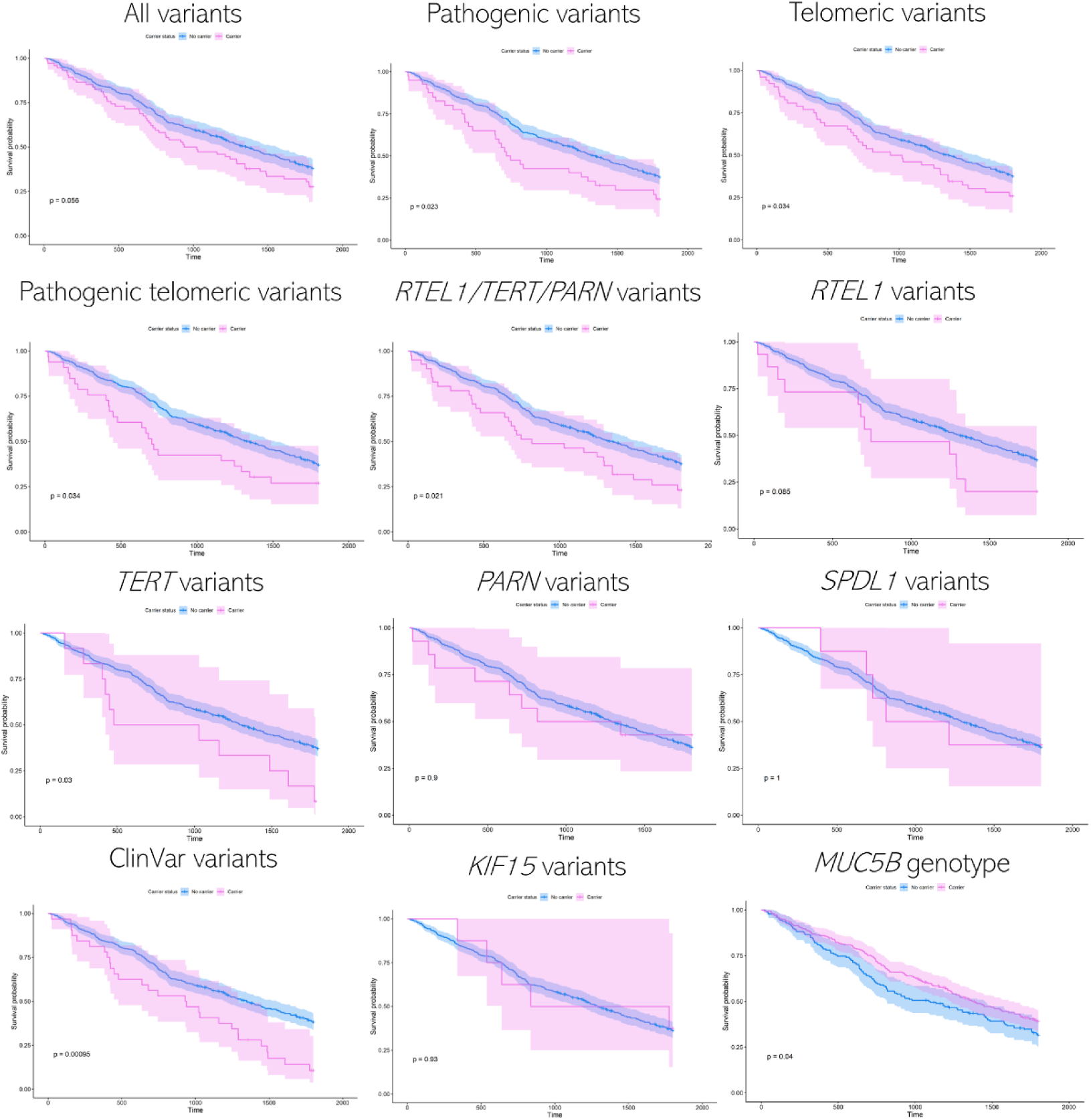
Kaplan-Meier survival analysis for qualifying variants (QV) (per gene and group PF genes) and the *MUC5B* risk allele in PROFILE. p-values for the log-rank test are shown.

